# REACT-1 round 12 report: resurgence of SARS-CoV-2 infections in England associated with increased frequency of the Delta variant

**DOI:** 10.1101/2021.06.17.21259103

**Authors:** Steven Riley, Haowei Wang, Oliver Eales, David Haw, Caroline E. Walters, Kylie E. C. Ainslie, Christina Atchison, Claudio Fronterre, Peter J. Diggle, Andrew J. Page, Sophie J. Prosolek, Alexander J. Trotter, Thanh Le Viet, Nabil-Fareed Alikhan, Leigh M Jackson, Catherine Ludden, The COVID-19 Genomics UK (COG-UK) Consortium, Deborah Ashby, Christl A. Donnelly, Graham Cooke, Wendy Barclay, Helen Ward, Ara Darzi, Paul Elliott

## Abstract

**Background:** England entered a third national lockdown from 6 January 2021 due to the COVID-19 pandemic. Despite a successful vaccine rollout during the first half of 2021, cases and hospitalisations have started to increase since the end of May as the SARS-CoV-2 Delta (B.1.617.2) variant increases in frequency. The final step of relaxation of COVID-19 restrictions in England has been delayed from 21 June to 19 July 2021.

**Methods:** The REal-time Assessment of Community Transmision-1 (REACT-1) study measures the prevalence of swab-positivity among random samples of the population of England. Round 12 of REACT-1 obtained self-administered swab collections from participants from 20 May 2021 to 7 June 2021; results are compared with those for round 11, in which swabs were collected from 15 April to 3 May 2021.

**Results:** Between rounds 11 and 12, national prevalence increased from 0.10% (0.08%, 0.13%) to 0.15% (0.12%, 0.18%). During round 12, we detected exponential growth with a doubling time of 11 (7.1, 23) days and an R number of 1.44 (1.20, 1.73). The highest prevalence was found in the North West at 0.26% (0.16%, 0.41%) compared to 0.05% (0.02%, 0.12%) in the South West. In the North West, the locations of positive samples suggested a cluster in Greater Manchester and the east Lancashire area. Prevalence in those aged 5-49 was 2.5 times higher at 0.20% (0.16%, 0.26%) compared with those aged 50 years and above at 0.08% (0.06%, 0.11%). At the beginning of February 2021, the link between infection rates and hospitalisations and deaths started to weaken, although in late April 2021, infection rates and hospital admissions started to reconverge. When split by age, the weakened link between infection rates and hospitalisations at ages 65 years and above was maintained, while the trends converged below the age of 65 years. The majority of the infections in the younger group occurred in the unvaccinated population or those without a stated vaccine history. We observed the rapid replacement of the Alpha (B.1.1.7) variant of SARS-CoV-2 with the Delta variant during the period covered by rounds 11 and 12 of the study.

**Discussion:** The extent to which exponential growth continues, or slows down as a consequence of the continued rapid roll-out of the vaccination programme, including to young adults, requires close monitoring. Data on community prevalence are vital to track the course of the epidemic and inform ongoing decisions about the timing of further lifting of restrictions in England.

## Introduction

The global distribution of COVID-19 cases and deaths is being driven by the emergence of more transmissible variants [1], variation in levels of population immunity obtained either from infection or vaccination [2], and by the degree of physical, social and workplace mixing [3]. Since late December 2020, in England, a successful vaccination campaign [4] has substantially increased population immunity while the gradual easing of the third national lockdown has led to an increase in social mixing [5]. Since the end of April 2021, the Delta (B.1.617.2) variant, first identified in India, has been replacing the Alpha (B.1.1.7) variant, first identified in the UK [6], in routinely collected genomic data [7], and case numbers and hospitalisations have started to rise [8]. These factors informed the decision to implement a four-week delay in the final stage of easing lockdown restrictions in England from 21 June to 19 July [9].

Here we report results from the twelfth round of the REal-time Assessment of Community Transmission-1 study (REACT-1) involving a random sample of the population of England. We invited named individuals to provide a throat and nose swab for RT-PCR testing for SARS-CoV-2 virus, and to answer an online questionnaire [10,11]. In round 12 we obtained self-administered swab collections from participants from 20 May 2021 to 7 June 2021. We compare these results to those from round 11 of REACT-1, in which swabs were collected from 15 April to 3 May 2021.

## Results

In round 12 we obtained 135 positives overall from 108,911 valid swabs giving a weighted prevalence of 0.15% (0.12%, 0.18%) (Table 1). This compares with weighted prevalence in round 11 of 0.10% (0.08%, 0.13%).

**Table 1.** The unweighted and weighted prevalence of swab-positivity across 12 rounds of REACT-1.

| Round | Tested swabs | Positive swabs | Unweighted prevalence (95% CI) | Weighted prevalence (95% CI) | First sample | Last sample |
| --- | --- | --- | --- | --- | --- | --- |
| 1 | 120,620 | 159 | 0.13% (0.11%, 0.15%) | 0.16% (0.13%, 0.19%) | 1/5/2020 | 1/6/2020 |
| 2 | 159,199 | 123 | 0.077% (0.065%, 0.092%) | 0.088% (0.068%, 0.11%) | 19/6/2020 | 7/7/2020 |
| 3 | 162,821 | 54 | 0.033% (0.025%, 0.043%) | 0.040% (0.027%, 0.053%) | 24/7/2020 | 11/8/2020 |
| 4 | 154,325 | 137 | 0.089% (0.075%, 0.11%) | 0.13% (0.096%, 0.15%) | 20/8/2020 | 8/9/2020 |
| 5 | 174,949 | 824 | 0.47% (0.44%, 0.50%) | 0.60% (0.55%, 0.71%) | 18/9/2020 | 5/10/2020 |
| 6 | 160,175 | 1,732 | 1.08% (1.03%, 1.13%) | 1.30% (1.21%, 1.39%) | 16/10/2020 | 2/11/2020 |
| 7 | 168,181 | 1,299 | 0.77% (0.73%, 0.82%) | 0.94% (0.87%, 1.01%) | 13/11/2020 | 3/12/2020 |
| 8 | 167,642 | 2,282 | 1.36% (1.31%, 1.42%) | 1.57% (1.49%, 1.66%) | 06/01/2021* | 22/01/2021 |
| 9 | 165,456 | 689 | 0.42% (0.39%, 0.45%) | 0.49% (0.44%, 0.55%) | 4/2/2021 | 23/2/2021 |
| 10 | 140,844 | 227 | 0.16% (0.14%, 0.18%) | 0.20% (0.17%, 0.23%) | 11/03/2021 | 30/3/2021 |
| 11 | 127,408 | 115 | 0.09% (0.07%, 0.11%) | 0.10% (0.08%, 0.13%) | 15/04/2021 | 3/5/2021 |
| 12 | 108,911 | 135 | 0.12% (0.10%, 0.15%) | 0.15% (0.12%, 0.18%) | 20/05/2021 | 07/06/2021 |

\* Includes small number of samples from previous days

We saw exponential growth in the proportion of swabs that were positive during round 12 (Figure 1). Using constant growth rate models we found strong evidence for a recent increase in R during round 12: R was 1.44 (1.20, 1.73) with >99% probability that R > 1 (Table 2) and with a doubling time of 11 (7.1, 23) days; the rate of growth was slightly lower with use of a P-spline model (Figure 2). From round 11 to round 12, R was 1.07 (1.03, 1.12). After previously documented declines in swab positivity in England [12] we estimate a turning point on or around 13 May (18 April, 21 May) after which prevalence started to increase (Figure 1, Figure 3).

**Table 2.** Estimates of national growth rates, doubling times and reproduction numbers for round 12, and round 11 to round 12.

| Round | Outcome | Growth rate | R | Probability R>1 | Doubling (+) / Halving (-) time |
| --- | --- | --- | --- | --- | --- |
| 12 | All positives | 0.063 ( 0.030 , 0.097 ) | 1.44 ( 1.20 , 1.73 ) | >0.99 | 11.1 ( 23.4 , 7.1 ) |
|  | Non-symptomatics | 0.030 ( -0.027 , 0.087 ) | 1.20 ( 0.84 , 1.64 ) | 0.85 | 23.3 ( -26.0 , 8.0 ) |
|  | Positive for both E and N genes | 0.104 ( 0.064 , 0.146 ) | 1.79 ( 1.45 , 2.18 ) | >0.99 | 6.7 ( 10.8 , 4.8 ) |
|  | Positive for both E and N genes or positive only for N gene with CT 35 or less | 0.073 ( 0.037 , 0.110 ) | 1.53 ( 1.25 , 1.84 ) | >0.99 | 9.5 ( 18.7 , 6.3 ) |
| 11 and 12 | All positives | 0.012 ( 0.005 , 0.018 ) | 1.07 ( 1.03 , 1.12 ) | >0.99 | * ( * , 38.4 ) |
|  | Non-symptomatics | 0.000 ( -0.010 , 0.010 ) | 1.00 ( 0.94 , 1.07 ) | 0.53 | * ( * , * ) |
|  | Positive for both E and N genes | 0.019 ( 0.011 , 0.027 ) | 1.13 ( 1.07 , 1.18 ) | >0.99 | 36.2 ( * , 25.3 ) |
|  | Positive for both E and N genes or positive only for N gene with CT 35 or less | 0.015 ( 0.008 , 0.022 ) | 1.10 ( 1.05 , 1.15 ) | >0.99 | 46.3 ( * , 31.3 ) |

\* Doubling/Halving time had an estimated magnitude greater than 50 days and so represented approximately constant prevalence

**Figure 1.**
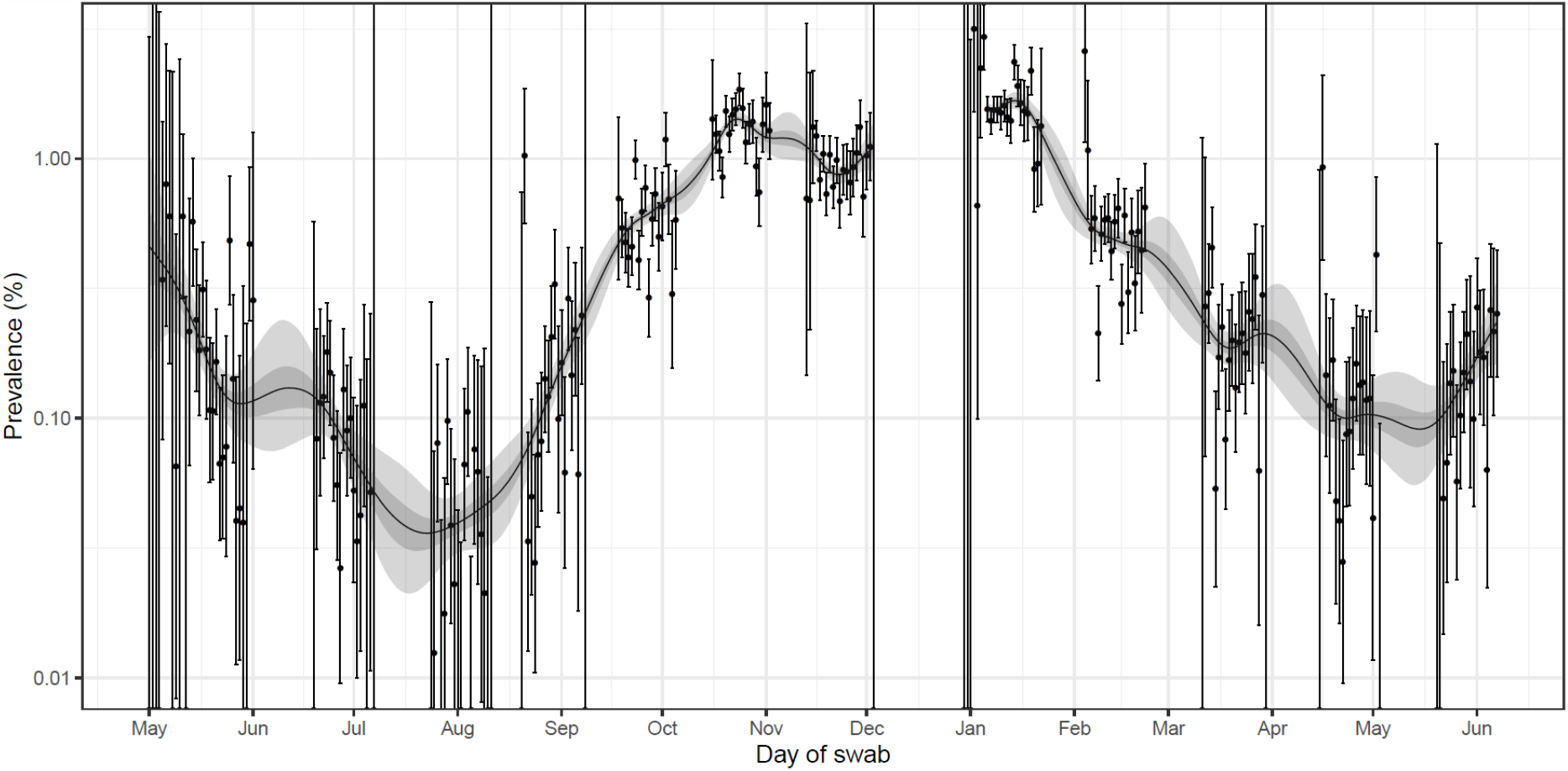
Prevalence of national swab-positivity for England estimated using a P-spline for all twelve rounds with central 50% (dark grey) and 95% (light grey) posterior credible intervals. Shown here for the entire period of the study with a log_10_ y-axis. Weighted observations (black dots) and 95% binomial confidence intervals (vertical lines) are also shown. Note that the period between round 7 and round 8 (December) of the model is not included as there were no data available to capture the late December peak of the epidemic.

**Figure 2.**
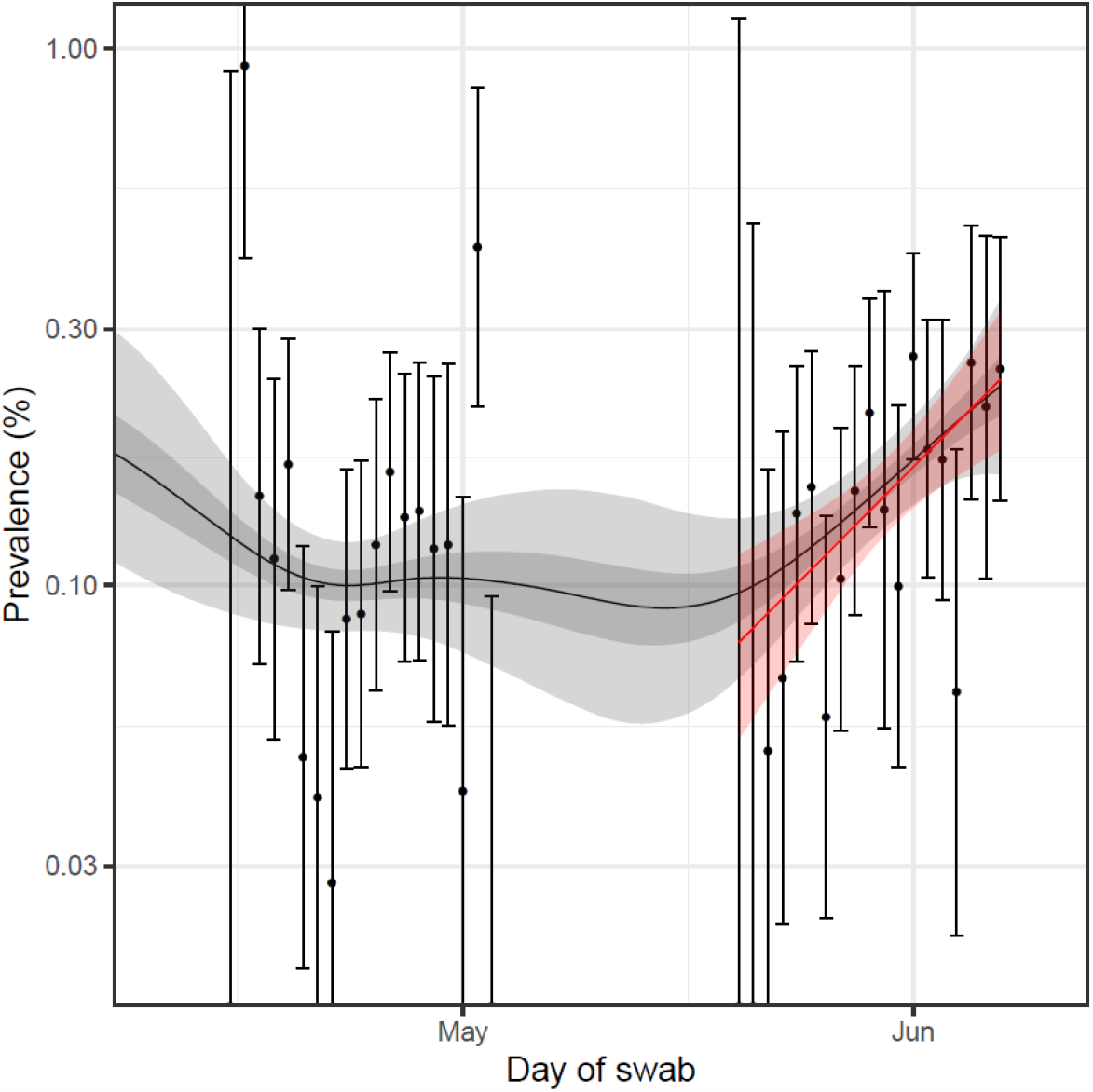
Comparison of the P-spline model and exponential model fit to round 12 only. Prevalence of national swab-positivity for England estimated using a P-spline for all twelve rounds with central 50% (dark grey) and 95% (light grey) posterior credible intervals. Shown here for rounds 11 and 12 with a log_10_ y-axis. Prevalence of national swab-positivity for England estimated using an exponential model for round 12 (red) and 95% credible intervals (light red). Weighted observations (black dots) and 95% binomial confidence intervals (vertical lines) are also shown.

**Figure 3.**
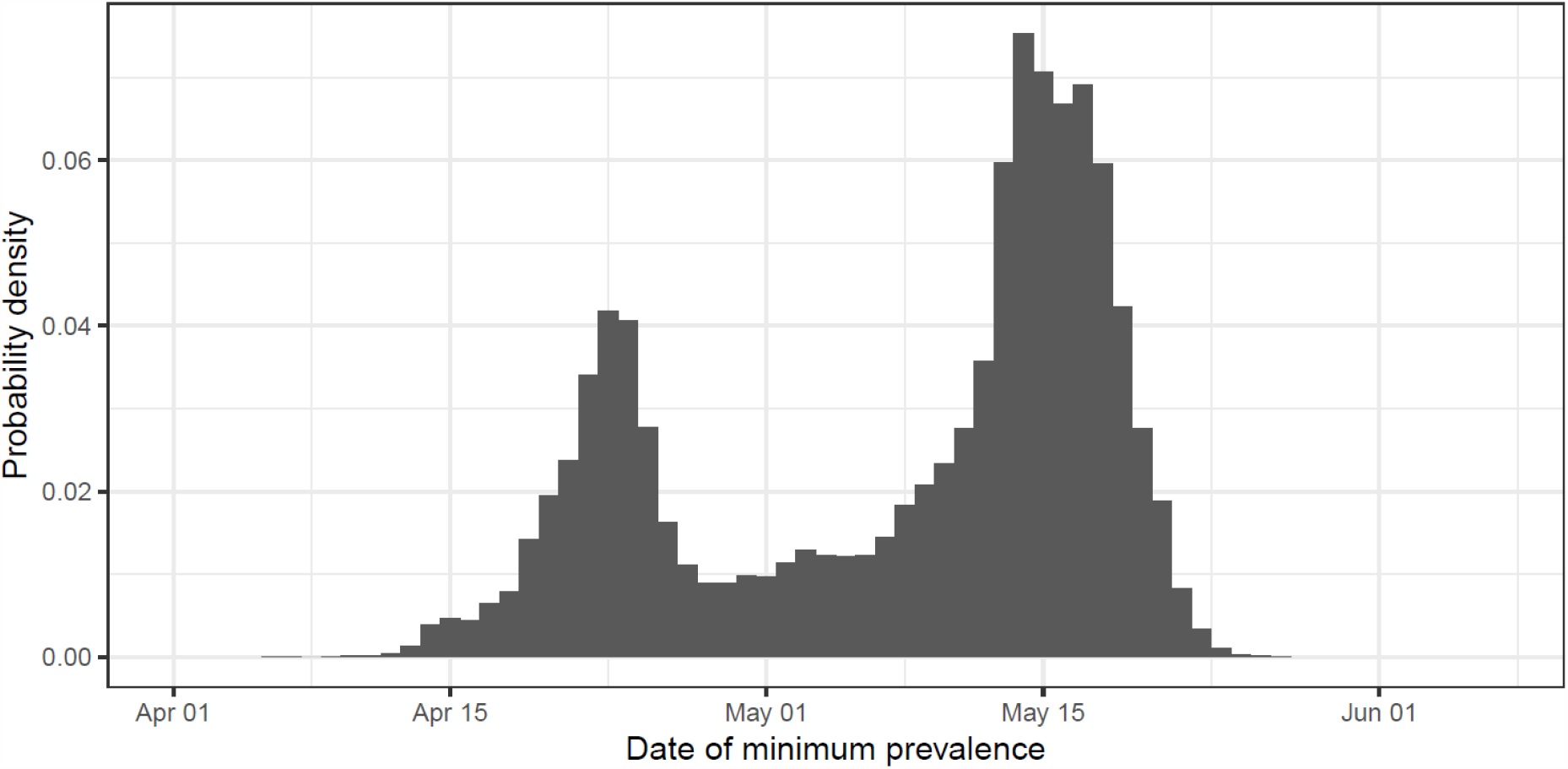
Probability density of the date of minimum prevalence as inferred from the national P-spline model.

Regional R between rounds 11 and 12 was above one with probability ≥95% in the North West, East Midlands and East of England (Table 3). For other regions, there was no strong evidence that R was different from one between rounds 11 and 12. There were too few data to estimate regional R within round 12 alone. We therefore estimated P-splines for the five northernmost regions combined and the four southernmost regions combined and found a sustained upward trend in the North with more recent growth in the South (Figure 4).

**Table 3.** Estimates of regional growth rates, doubling times and reproduction numbers for round 11 to round 12.

| Round | Region | Growth rate | R | Probability R>1 | Halving (-) / Doubling (+) time |
| --- | --- | --- | --- | --- | --- |
| 11 and 12 | East Midlands | 0.028 ( 0.006 , 0.052 ) | 1.19 ( 1.04 , 1.37 ) | 0.99 | 24.8 ( * , 13.2 ) |
|  | West Midlands | -0.002 ( -0.023 , 0.018 ) | 0.99 ( 0.86 , 1.12 ) | 0.42 | * ( -30.3 , 38.6 ) |
|  | East of England | 0.018 ( -0.003 , 0.040 ) | 1.12 ( 0.98 , 1.27 ) | 0.95 | 39.0 ( * , 17.5 ) |
|  | London | 0.002 ( -0.015 , 0.020 ) | 1.02 ( 0.91 , 1.13 ) | 0.61 | * ( -47.5 , 35.4 ) |
|  | North West | 0.022 ( 0.007 , 0.039 ) | 1.15 ( 1.04 , 1.26 ) | >0.99 | 31.4 ( * , 17.9 ) |
|  | North East | 0.009 ( -0.022 , 0.040 ) | 1.06 ( 0.87 , 1.27 ) | 0.72 | * ( -32.2 , 17.3 ) |
|  | South East | 0.008 ( -0.007 , 0.024 ) | 1.05 ( 0.95 , 1.16 ) | 0.85 | * ( * , 28.4 ) |
|  | South West | -0.004 ( -0.035 , 0.025 ) | 0.97 ( 0.79 , 1.17 ) | 0.39 | * ( -19.8 , 27.8 ) |
|  | Yorkshire ** | 0.016 ( -0.004 , 0.036 ) | 1.10 ( 0.98 , 1.24 ) | 0.94 | 44.3 ( * , 19.2 ) |

\* Doubling/Halving time had an estimated magnitude greater than 50 days and so represented approximately constant prevalence
\*\* Yorkshire and The Humber

**Figure 4.**
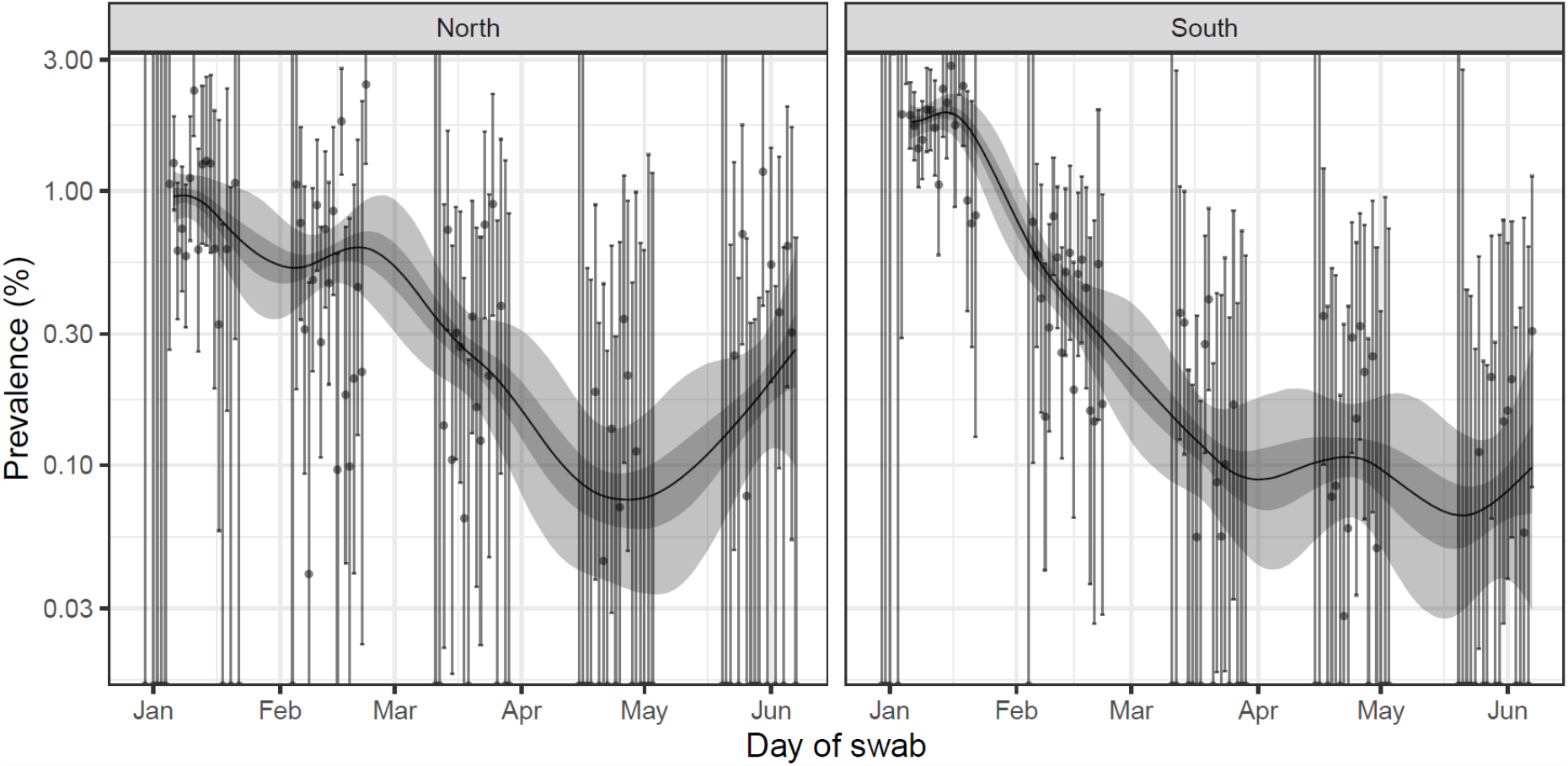
Prevalence of swab-positivity for the North of England (North West, North East, Yorkshire and The Humber, East Midlands and West Midlands) and the South of England (East of England, London, South East and South West) estimated using a P-spline for all twelve rounds with central 50% (dark grey) and 95% (light grey) posterior credible intervals. Shown here for the period of the study since January 2021 with a log_10_ y-axis. Weighted observations (black dots) and 95% binomial confidence intervals (vertical lines) are also shown.

We found substantial heterogeneity in prevalence between regions. Weighted prevalence was higher in the North West at 0.26% (0.16%, 0.41%) compared to 0.05% (0.02%, 0.12%) in the South West (Table 4, Figure 5, Figure 6). In the North West, the locations of positive samples suggested a cluster in Greater Manchester and the east Lancashire area.

**Table 4a.**
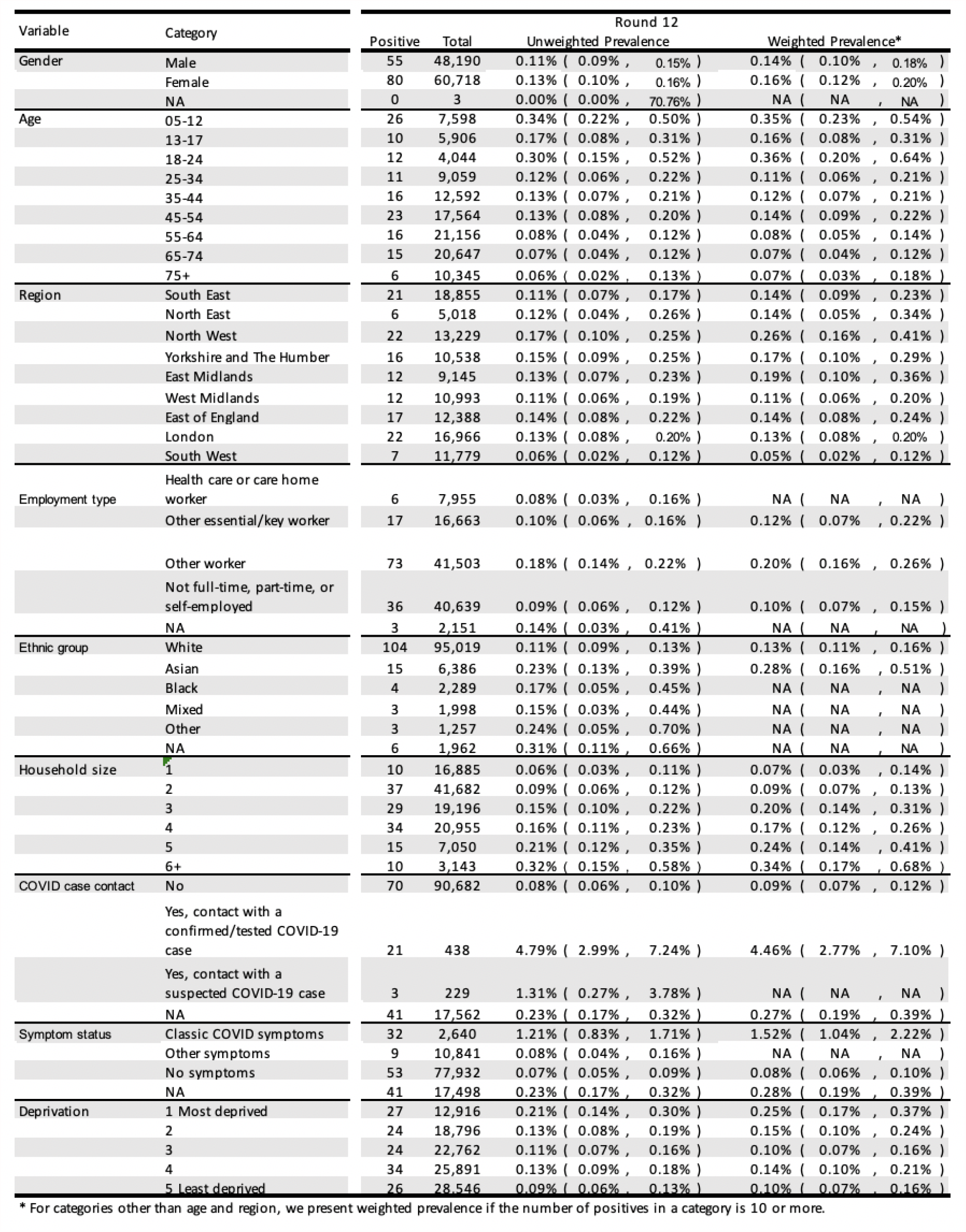
Unweighted and weighted prevalence of swab-positivity for sex, age, region and other key variables for round 12.

| Variable | Category | Round 12 |  |  |  |
| --- | --- | --- | --- | --- | --- |
|  |  | Positive | Total | Unweighted Prevalence | Weighted Prevalence* |
| Gender | Male | 55 | 48,190 | 0.11% ( 0.09% , 0.15% ) | 0.14% ( 0.10% , 0.18% ) |
|  | Female | 80 | 60,718 | 0.13% ( 0.10% , 0.16% ) | 0.16% ( 0.12% , 0.20% ) |
|  | NA | 0 | 3 | 0.00% ( 0.00% , 70.76% ) | NA ( NA , NA ) |
| Age | 05-12 | 26 | 7,598 | 0.34% ( 0.22% , 0.50% ) | 0.35% ( 0.23% , 0.54% ) |
|  | 13-17 | 10 | 5,906 | 0.17% ( 0.08% , 0.31% ) | 0.16% ( 0.08% , 0.31% ) |
|  | 18-24 | 12 | 4,044 | 0.30% ( 0.15% , 0.52% ) | 0.36% ( 0.20% , 0.64% ) |
|  | 25-34 | 11 | 9,059 | 0.12% ( 0.06% , 0.22% ) | 0.11% ( 0.06% , 0.21% ) |
|  | 35-44 | 16 | 12,592 | 0.13% ( 0.07% , 0.21% ) | 0.12% ( 0.07% , 0.21% ) |
|  | 45-54 | 23 | 17,564 | 0.13% ( 0.08% , 0.20% ) | 0.14% ( 0.09% , 0.22% ) |
|  | 55-64 | 16 | 21,156 | 0.08% ( 0.04% , 0.12% ) | 0.08% ( 0.05% , 0.14% ) |
|  | 65-74 | 15 | 20,647 | 0.07% ( 0.04% , 0.12% ) | 0.07% ( 0.04% , 0.12% ) |
|  | 75+ | 6 | 10,345 | 0.06% ( 0.02% , 0.13% ) | 0.07% ( 0.03% , 0.18% ) |
| Region | South East | 21 | 18,855 | 0.11% ( 0.07% , 0.17% ) | 0.14% ( 0.09% , 0.23% ) |
|  | North East | 6 | 5,018 | 0.12% ( 0.04% , 0.26% ) | 0.14% ( 0.05% , 0.34% ) |
|  | North West | 22 | 13,229 | 0.17% ( 0.10% , 0.25% ) | 0.26% ( 0.16% , 0.41% ) |
|  | Yorkshire and The Humber | 16 | 10,538 | 0.15% ( 0.09% , 0.25% ) | 0.17% ( 0.10% , 0.29% ) |
|  | East Midlands | 12 | 9,145 | 0.13% ( 0.07% , 0.23% ) | 0.19% ( 0.10% , 0.36% ) |
|  | West Midlands | 12 | 10,993 | 0.11% ( 0.06% , 0.19% ) | 0.11% ( 0.06% , 0.20% ) |
|  | East of England | 17 | 12,388 | 0.14% ( 0.08% , 0.22% ) | 0.14% ( 0.08% , 0.24% ) |
|  | London | 22 | 16,966 | 0.13% ( 0.08% , 0.20% ) | 0.13% ( 0.08% , 0.20% ) |
|  | South West | 7 | 11,779 | 0.06% ( 0.02% , 0.12% ) | 0.05% ( 0.02% , 0.12% ) |
| Employment type | Health care or care home worker | 6 | 7,955 | 0.08% ( 0.03% , 0.16% ) | NA ( NA , NA ) |
|  | Other essential/key worker | 17 | 16,663 | 0.10% ( 0.06% , 0.16% ) | 0.12% ( 0.07% , 0.22% ) |
|  | Other worker | 73 | 41,503 | 0.18% ( 0.14% , 0.22% ) | 0.20% ( 0.16% , 0.26% ) |
|  | Not full-time, part-time, or self-employed | 36 | 40,639 | 0.09% ( 0.06% , 0.12% ) | 0.10% ( 0.07% , 0.15% ) |
|  | NA | 3 | 2,151 | 0.14% ( 0.03% , 0.41% ) | NA ( NA , NA ) |
| Ethnic group | White | 104 | 95,019 | 0.11% ( 0.09% , 0.13% ) | 0.13% ( 0.11% , 0.16% ) |
|  | Asian | 15 | 6,386 | 0.23% ( 0.13% , 0.39% ) | 0.28% ( 0.16% , 0.51% ) |
|  | Black | 4 | 2,289 | 0.17% ( 0.05% , 0.45% ) | NA ( NA , NA ) |
|  | Mixed | 3 | 1,998 | 0.15% ( 0.03% , 0.44% ) | NA ( NA , NA ) |
|  | Other | 3 | 1,257 | 0.24% ( 0.05% , 0.70% ) | NA ( NA , NA ) |
|  | NA | 6 | 1,962 | 0.31% ( 0.11% , 0.66% ) | NA ( NA , NA ) |
| Household size | 1 | 10 | 16,885 | 0.06% ( 0.03% , 0.11% ) | 0.07% ( 0.03% , 0.14% ) |
|  | 2 | 37 | 41,682 | 0.09% ( 0.06% , 0.12% ) | 0.09% ( 0.07% , 0.13% ) |
|  | 3 | 29 | 19,196 | 0.15% ( 0.10% , 0.22% ) | 0.20% ( 0.14% , 0.31% ) |
|  | 4 | 34 | 20,955 | 0.16% ( 0.11% , 0.23% ) | 0.17% ( 0.12% , 0.26% ) |
|  | 5 | 15 | 7,050 | 0.21% ( 0.12% , 0.35% ) | 0.24% ( 0.14% , 0.41% ) |
|  | 6+ | 10 | 3,143 | 0.32% ( 0.15% , 0.58% ) | 0.34% ( 0.17% , 0.68% ) |
| COVID case contact | No | 70 | 90,682 | 0.08% ( 0.06% , 0.10% ) | 0.09% ( 0.07% , 0.12% ) |
|  | Yes, contact with a confirmed/tested COVID-19 case | 21 | 438 | 4.79% ( 2.99% , 7.24% ) | 4.46% ( 2.77% , 7.10% ) |
|  | Yes, contact with a suspected COVID-19 case | 3 | 229 | 1.31% ( 0.27% , 3.78% ) | NA ( NA , NA ) |
|  | NA | 41 | 17,562 | 0.23% ( 0.17% , 0.32% ) | 0.27% ( 0.19% , 0.39% ) |
| Symptom status | Classic COVID symptoms | 32 | 2,640 | 1.21% ( 0.83% , 1.71% ) | 1.52% ( 1.04% , 2.22% ) |
|  | Other symptoms | 9 | 10,841 | 0.08% ( 0.04% , 0.16% ) | NA ( NA , NA ) |
|  | No symptoms | 53 | 77,932 | 0.07% ( 0.05% , 0.09% ) | 0.08% ( 0.06% , 0.10% ) |
|  | NA | 41 | 17,498 | 0.23% ( 0.17% , 0.32% ) | 0.28% ( 0.19% , 0.39% ) |
| Deprivation | 1 Most deprived | 27 | 12,916 | 0.21% ( 0.14% , 0.30% ) | 0.25% ( 0.17% , 0.37% ) |
|  | 2 | 24 | 18,796 | 0.13% ( 0.08% , 0.19% ) | 0.15% ( 0.10% , 0.24% ) |
|  | 3 | 24 | 22,762 | 0.11% ( 0.07% , 0.16% ) | 0.10% ( 0.07% , 0.16% ) |
|  | 4 | 34 | 25,891 | 0.13% ( 0.09% , 0.18% ) | 0.14% ( 0.10% , 0.21% ) |
|  | 5 Least deprived | 26 | 28,546 | 0.09% ( 0.06% , 0.13% ) | 0.10% ( 0.07% , 0.16% ) |

\* For categories other than age and region, we present weighted prevalence if the number of positives in a category is 10 or more.

**Table 4b.**
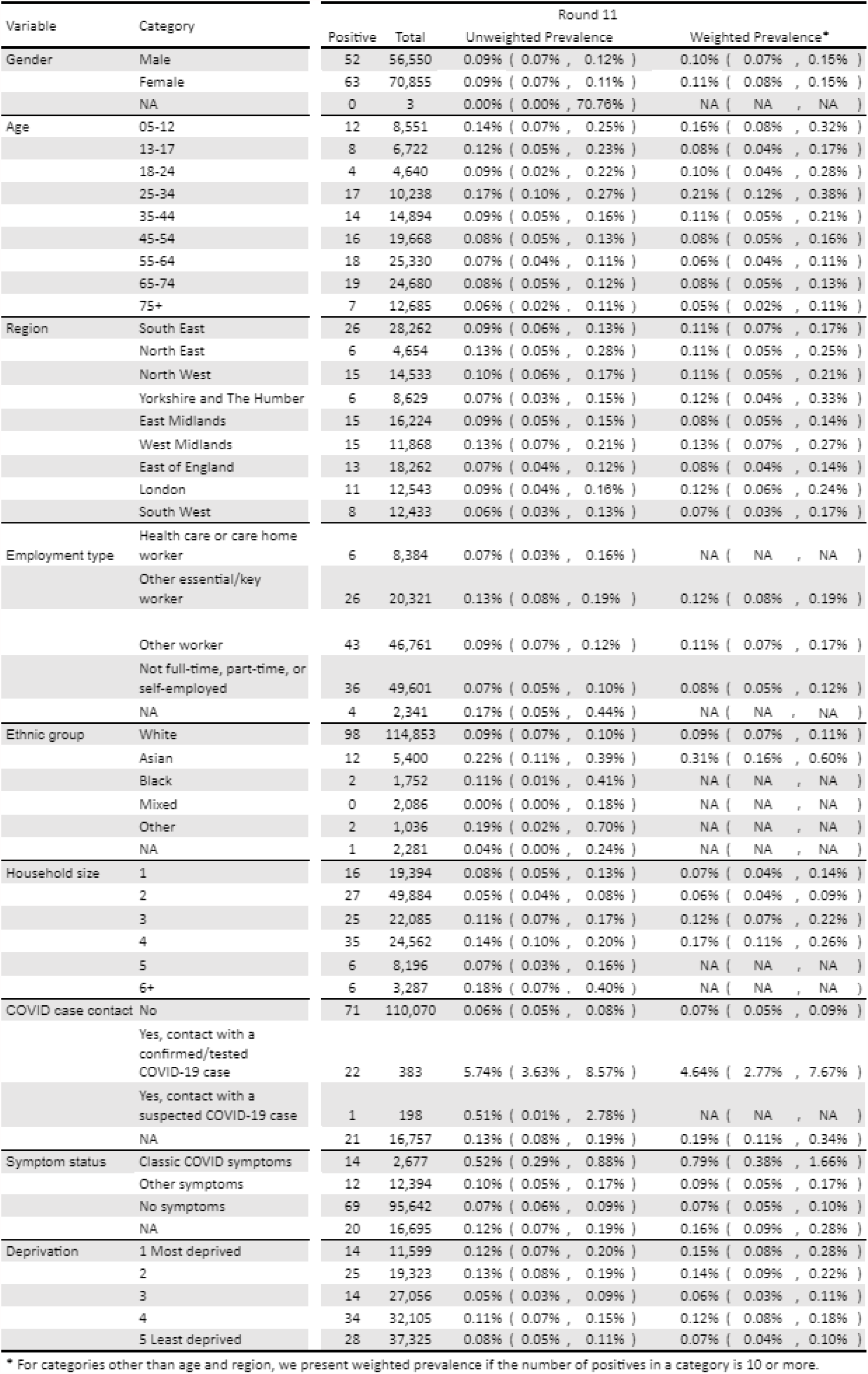
Unweighted and weighted prevalence of swab-positivity for sex, age, region and other key variables for round 11.

| Variable | Category | Round 11 |  |  |  |
| --- | --- | --- | --- | --- | --- |
|  |  | Positive | Total | Unweighted Prevalence | Weighted Prevalence* |
| Gender | Male | 52 | 56,550 | 0.09% ( 0.07% , 0.12% ) | 0.10% ( 0.07% , 0.15% ) |
|  | Female | 63 | 70,855 | 0.09% ( 0.07% , 0.11% ) | 0.11% ( 0.08% , 0.15% ) |
|  | NA | 0 | 3 | 0.00% ( 0.00% , 70.76% ) | NA ( NA , NA ) |
| Age | 05-12 | 12 | 8,551 | 0.14% ( 0.07% , 0.25% ) | 0.16% ( 0.08% , 0.32% ) |
|  | 13-17 | 8 | 6,722 | 0.12% ( 0.05% , 0.23% ) | 0.08% ( 0.04% , 0.17% ) |
|  | 18-24 | 4 | 4,640 | 0.09% ( 0.02% , 0.22% ) | 0.10% ( 0.04% , 0.28% ) |
|  | 25-34 | 17 | 10,238 | 0.17% ( 0.10% , 0.27% ) | 0.21% ( 0.12% , 0.38% ) |
|  | 35-44 | 14 | 14,894 | 0.09% ( 0.05% , 0.16% ) | 0.11% ( 0.05% , 0.21% ) |
|  | 45-54 | 16 | 19,668 | 0.08% ( 0.05% , 0.13% ) | 0.08% ( 0.05% , 0.16% ) |
|  | 55-64 | 18 | 25,330 | 0.07% ( 0.04% , 0.11% ) | 0.06% ( 0.04% , 0.11% ) |
|  | 65-74 | 19 | 24,680 | 0.08% ( 0.05% , 0.12% ) | 0.08% ( 0.05% , 0.13% ) |
|  | 75+ | 7 | 12,685 | 0.06% ( 0.02% , 0.11% ) | 0.05% ( 0.02% , 0.11% ) |
| Region | South East | 26 | 28,262 | 0.09% ( 0.06% , 0.13% ) | 0.11% ( 0.07% , 0.17% ) |
|  | North East | 6 | 4,654 | 0.13% ( 0.05% , 0.28% ) | 0.11% ( 0.05% , 0.25% ) |
|  | North West | 15 | 14,533 | 0.10% ( 0.06% , 0.17% ) | 0.11% ( 0.05% , 0.21% ) |
|  | Yorkshire and The Humber | 6 | 8,629 | 0.07% ( 0.03% , 0.15% ) | 0.12% ( 0.04% , 0.33% ) |
|  | East Midlands | 15 | 16,224 | 0.09% ( 0.05% , 0.15% ) | 0.08% ( 0.05% , 0.14% ) |
|  | West Midlands | 15 | 11,868 | 0.13% ( 0.07% , 0.21% ) | 0.13% ( 0.07% , 0.27% ) |
|  | East of England | 13 | 18,262 | 0.07% ( 0.04% , 0.12% ) | 0.08% ( 0.04% , 0.14% ) |
|  | London | 11 | 12,543 | 0.09% ( 0.04% , 0.16% ) | 0.12% ( 0.06% , 0.24% ) |
|  | South West | 8 | 12,433 | 0.06% ( 0.03% , 0.13% ) | 0.07% ( 0.03% , 0.17% ) |
| Employment type | Health care or care home worker | 6 | 8,384 | 0.07% ( 0.03% , 0.16% ) | NA ( NA , NA ) |
|  | Other essential/key worker | 26 | 20,321 | 0.13% ( 0.08% , 0.19% ) | 0.12% ( 0.08% , 0.19% ) |
|  | Other worker | 43 | 46,761 | 0.09% ( 0.07% , 0.12% ) | 0.11% ( 0.07% , 0.17% ) |
|  | Not full-time, part-time, or self-employed | 36 | 49,601 | 0.07% ( 0.05% , 0.10% ) | 0.08% ( 0.05% , 0.12% ) |
|  | NA | 4 | 2,341 | 0.17% ( 0.05% , 0.44% ) | NA ( NA , NA ) |
| Ethnic group | White | 98 | 114,853 | 0.09% ( 0.07% , 0.10% ) | 0.09% ( 0.07% , 0.11% ) |
|  | Asian | 12 | 5,400 | 0.22% ( 0.11% , 0.39% ) | 0.31% ( 0.16% , 0.60% ) |
|  | Black | 2 | 1,752 | 0.11% ( 0.01% , 0.41% ) | NA ( NA , NA ) |
|  | Mixed | 0 | 2,086 | 0.00% ( 0.00% , 0.18% ) | NA ( NA , NA ) |
|  | Other | 2 | 1,036 | 0.19% ( 0.02% , 0.70% ) | NA ( NA , NA ) |
|  | NA | 1 | 2,281 | 0.04% ( 0.00% , 0.24% ) | NA ( NA , NA ) |
| Household size | 1 | 16 | 19,394 | 0.08% ( 0.05% , 0.13% ) | 0.07% ( 0.04% , 0.14% ) |
|  | 2 | 27 | 49,884 | 0.05% ( 0.04% , 0.08% ) | 0.06% ( 0.04% , 0.09% ) |
|  | 3 | 25 | 22,085 | 0.11% ( 0.07% , 0.17% ) | 0.12% ( 0.07% , 0.22% ) |
|  | 4 | 35 | 24,562 | 0.14% ( 0.10% , 0.20% ) | 0.17% ( 0.11% , 0.26% ) |
|  | 5 | 6 | 8,196 | 0.07% ( 0.03% , 0.16% ) | NA ( NA , NA ) |
|  | 6+ | 6 | 3,287 | 0.18% ( 0.07% , 0.40% ) | NA ( NA , NA ) |
| COVID case contact | No | 71 | 110,070 | 0.06% ( 0.05% , 0.08% ) | 0.07% ( 0.05% , 0.09% ) |
|  | Yes, contact with a confirmed/tested COVID-19 case | 22 | 383 | 5.74% ( 3.63% , 8.57% ) | 4.64% ( 2.77% , 7.67% ) |
|  | Yes, contact with a suspected COVID-19 case | 1 | 198 | 0.51% ( 0.01% , 2.78% ) | NA ( NA , NA ) |
|  | NA | 21 | 16,757 | 0.13% ( 0.08% , 0.19% ) | 0.19% ( 0.11% , 0.34% ) |
| Symptom status | Classic COVID symptoms | 14 | 2,677 | 0.52% ( 0.29% , 0.88% ) | 0.79% ( 0.38% , 1.66% ) |
|  | Other symptoms | 12 | 12,394 | 0.10% ( 0.05% , 0.17% ) | 0.09% ( 0.05% , 0.17% ) |
|  | No symptoms | 69 | 95,642 | 0.07% ( 0.06% , 0.09% ) | 0.07% ( 0.05% , 0.10% ) |
|  | NA | 20 | 16,695 | 0.12% ( 0.07% , 0.19% ) | 0.16% ( 0.09% , 0.28% ) |
| Deprivation | 1 Most deprived | 14 | 11,599 | 0.12% ( 0.07% , 0.20% ) | 0.15% ( 0.08% , 0.28% ) |
|  | 2 | 25 | 19,323 | 0.13% ( 0.08% , 0.19% ) | 0.14% ( 0.09% , 0.22% ) |
|  | 3 | 14 | 27,056 | 0.05% ( 0.03% , 0.09% ) | 0.06% ( 0.03% , 0.11% ) |
|  | 4 | 34 | 32,105 | 0.11% ( 0.07% , 0.15% ) | 0.12% ( 0.08% , 0.18% ) |
|  | 5 Least deprived | 28 | 37,325 | 0.08% ( 0.05% , 0.11% ) | 0.07% ( 0.04% , 0.10% ) |

\* For categories other than age and region, we present weighted prevalence if the number of positives in a category is 10 or more.

**Figure 5.**
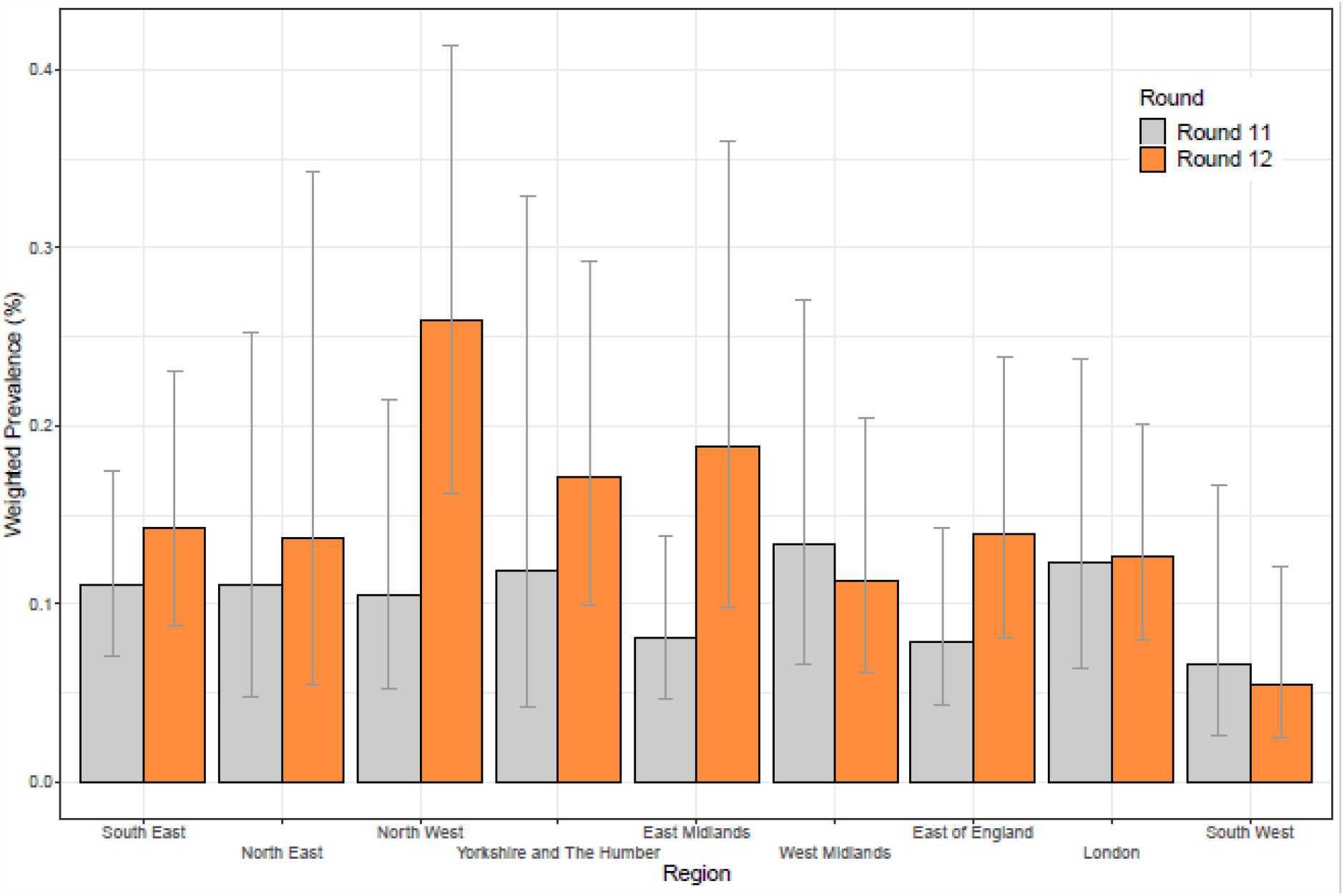
Weighted prevalence of swab-positivity by region for rounds 11 and 12. Bars show 95% confidence intervals.

**Figure 6.**
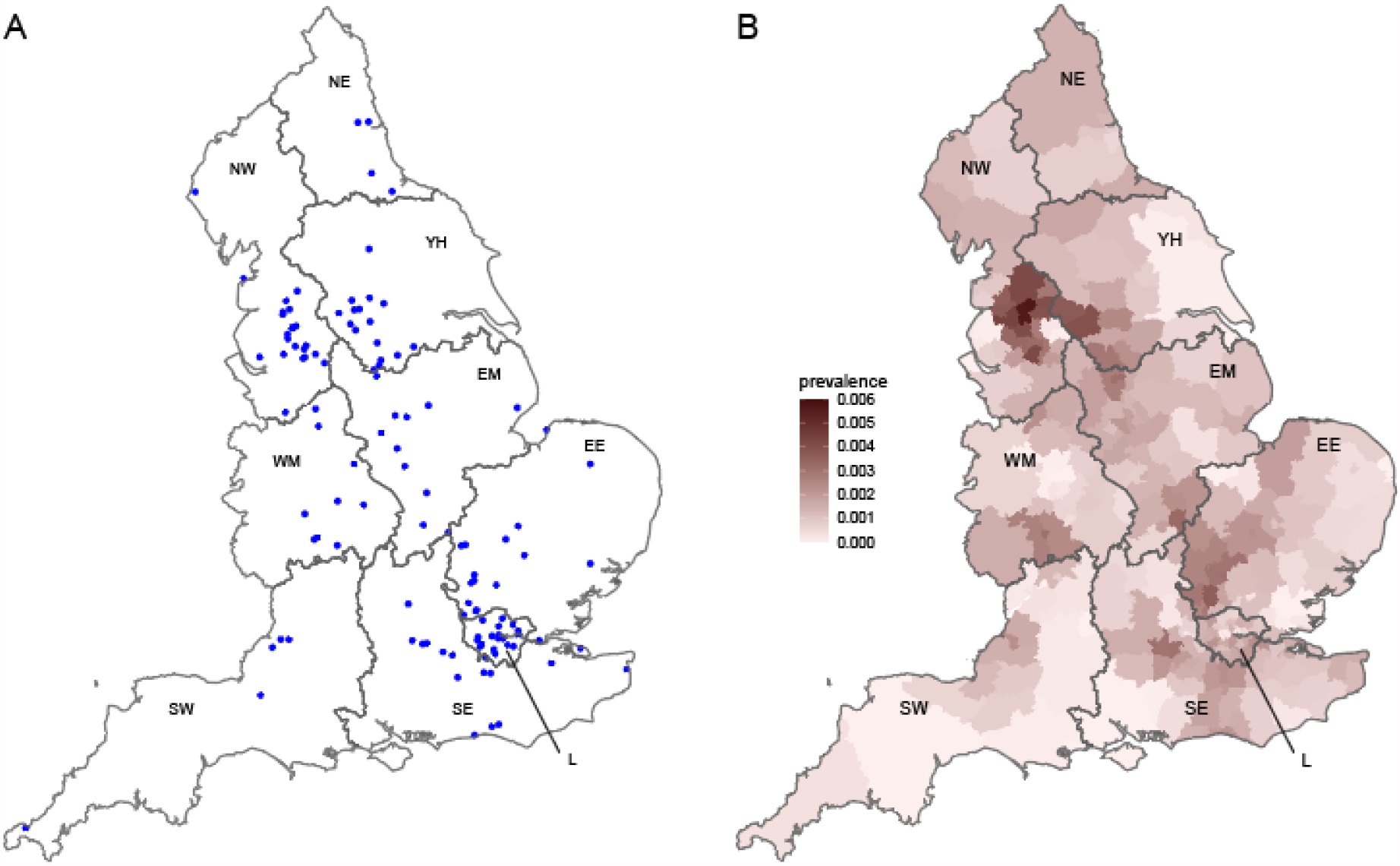
Geographical distribution of swab-positives in round 12. **A** Jittered location of all 135 positive samples detected. **B** Neighbourhood smoothed average prevalence by lower tier local area. Neighbourhood prevalence calculated from nearest neighbours (the median number of neighbours within 20 km in the study). Average neighbourhood prevalence displayed for individual lower-tier local authorities. Regions: NE = North East, NW = North West, YH = Yorkshire and The Humber, EM = East Midlands, WM = West Midlands, EE = East of England, L = London, SE = South East, SW = South West.

Weighted prevalence in round 12 was five-fold higher in 5-12 and 18-24 year olds at 0.35% (0.23%, 0.54%) and 0.36% (0.20%, 0.64%) respectively compared with people ages 65 and above (Table 4, Figure 7); and 2.5 times higher in those aged 5-49 years at 0.20% (0.16%, 0.26%) compared with those aged 50 years and above at 0.08% (0.06%, 0.11%) (Table 5). P-spline estimates for those aged 5-49 years and those aged 50 years and above showed similar recent upturns in prevalence, but at a higher level in the younger age group (Figure 8).

**Table 5.** Weighted prevalence of swab-positivity for 5-49 years and 50 years and above age group for rounds 11 and 12.

| Round | Category | Positive | Total | Weighted Prevalence |
| --- | --- | --- | --- | --- |
| 11 | 5-49 | 61 | 53,941 | 0.13% ( 0.10% , 0.18% ) |
|  | 50+ | 54 | 73,467 | 0.07% ( 0.05% , 0.09% ) |
| 12 | 5-49 | 86 | 47,053 | 0.20% ( 0.16% , 0.26% ) |
|  | 50+ | 49 | 61,858 | 0.08% ( 0.06% , 0.11% ) |

**Figure 7.**
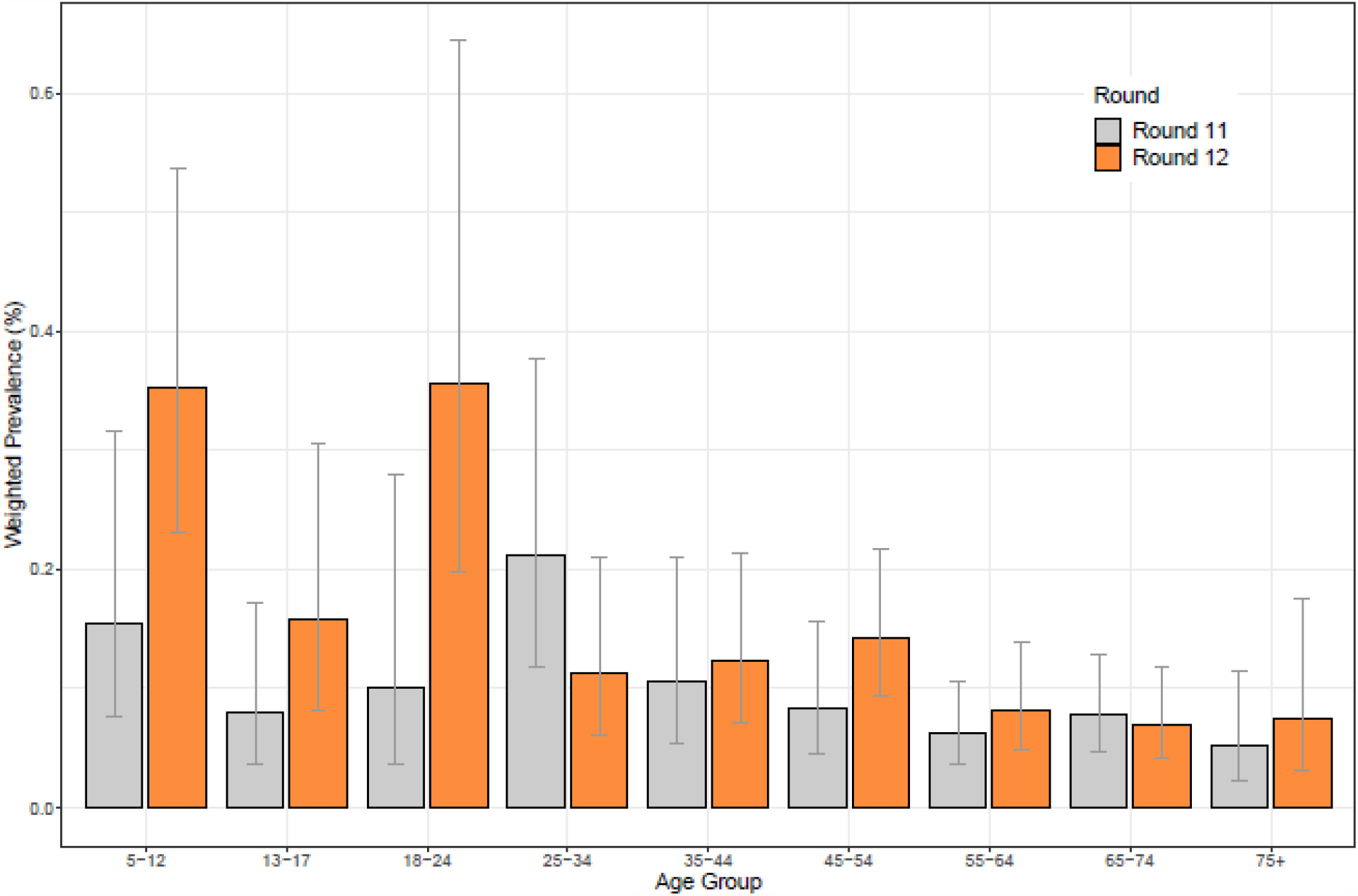
Weighted prevalence of swab-positivity by age for rounds 11 and 12. Bars show 95% confidence intervals.

**Figure 8.**
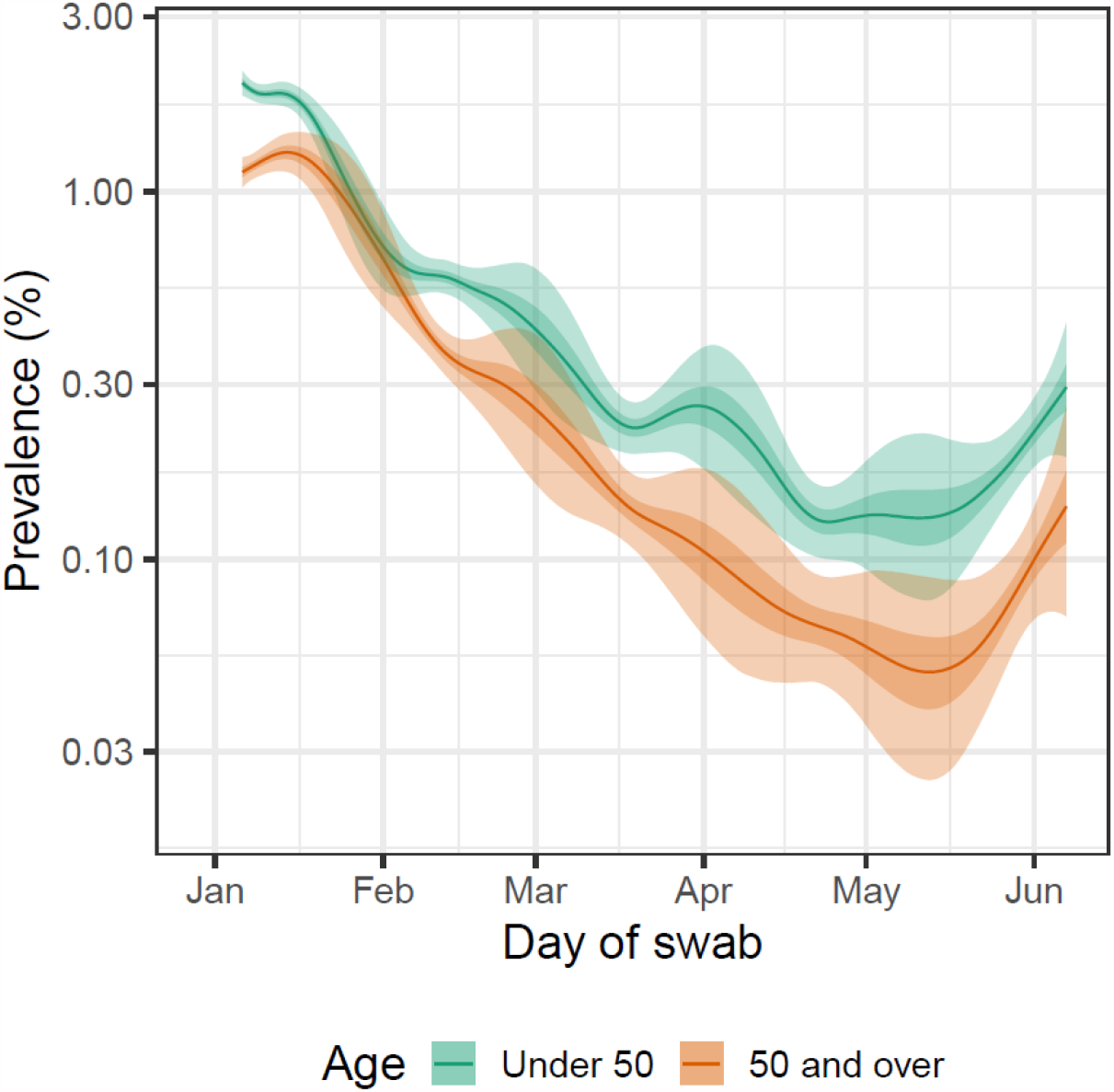
Prevalence of swab-positivity for those aged under 50 and those aged 50 and over estimated using a P-spline for all twelve rounds with central 50% (dark grey) and 95% (light grey) posterior credible intervals. Shown here for the period of the study since January with a log_10_ y-axis.

We carried out multivariable (mutually adjusted) logistic regression for key variables (Table 6). Odds of swab-positivity among healthcare and care home workers were reduced at 0.37 (0.16, 0.87) and other key workers at 0.55 (0.32,0.94) compared with other workers. Those living in the most deprived neighbourhoods had odds of swab-positivity of 1.94 (1.08, 3.51) compared with those living in the least deprived neighbourhoods.

**Table 6.** Multivariable logistic regression for rounds 11 and round 12.

| Variable | Category | Round 11* | Round 12* |
| --- | --- | --- | --- |
| Sex | Male | Ref | Ref |
|  | Female | 1.02 [0.69,1.49] | 1.34 [0.93,1.92] |
| Age Group | 5-12 | 1.52 [0.69,3.36] | 2.61 [1.32,5.17] |
|  | 13-17 | 1.66 [0.61,4.50] | 1.87 [0.76,4.63] |
|  | 18-24 | 1.03 [0.33,3.19] | 2.57 [1.18,5.64] |
|  | 25-34 | 2.02 [0.97,4.22] | 1.12 [0.50,2.47] |
|  | 35-44 | Ref | Ref |
|  | 45-54 | 1.01 [0.48,2.10] | 1.22 [0.62,2.39] |
|  | 55-64 | 1.09 [0.52,2.31] | 0.82 [0.39,1.72] |
|  | 65+ | 1.49 [0.66,3.33] | 0.80 [0.36,1.75] |
| Region | North East | 1.38 [0.56,3.42] | 1.14 [0.45,2.86] |
|  | North West | 1.10 [0.57,2.12] | 1.40 [0.75,2.63] |
|  | Yorkshire and The Hurn | 0.79 [0.32,1.95] | 1.39 [0.71,2.70] |
|  | East Midlands | 0.96 [0.50,1.86] | 1.20 [0.58,2.47] |
|  | West Midlands | 1.38 [0.72,2.63] | 0.77 [0.36,1.66] |
|  | East of England | 0.69 [0.34,1.41] | 1.06 [0.54,2.11] |
|  | London | 0.71 [0.33,1.53] | 0.84 [0.44,1.60] |
|  | South East | Ref | Ref |
|  | South West | 0.76 [0.34,1.69] | 0.58 [0.25,1.38] |
| Key Worker Status | HCW/CHW | 0.70 [0.29,1.66] | 0.37 [0.16,0.87] |
|  | Key worker (other) | 1.31 [0.79,2.15] | 0.55 [0.32,0.94] |
|  | Other worker | Ref | Ref |
|  | Not FT, PT, SE | 0.78 [0.46,1.31] | 0.60 [0.38,0.95] |
| Ethnicity | White | Ref | Ref |
|  | Asian | 1.95 [0.98,3.88] | 1.52 [0.84,2.76] |
|  | Black | 1.26 [0.30,5.25] | 1.26 [0.45,3.55] |
|  | Mixed | 0.00 [0.00,2.821236e+136] | 0.95 [0.30,3.05] |
|  | Other | 2.24 [0.54,9.27] | 1.82 [0.57,5.87] |
| Household Size | 1-2 People | Ref | Ref |
|  | 3-5 People | 1.84 [1.13,2.98] | 1.19 [0.75,1.89] |
|  | 6+ People | 2.19 [0.81,5.94] | 1.74 [0.78,3.88] |
| Deprivation Index Quintile | 1 - Most Deprived | 1.47 [0.75,2.88] | 1.94 [1.08,3.51] |
|  | 2 | 1.51 [0.85,2.69] | 1.39 [0.78,2.51] |
|  | 3 | 0.73 [0.38,1.40] | 1.11 [0.62,2.01] |
|  | 4 | 1.43 [0.86,2.39] | 1.55 [0.91,2.63] |
|  | 5 - Least Deprived | Ref | Ref |

\* Odds ratios mutually adjusted for all variables shown.

We investigated the relationship between swab-positivity, as estimated in rounds 1 to 11 of REACT-1, and deaths and hospital admissions with lags of 26 and 19 days respectively for all ages combined (Figure 9). Since the beginning of February the trend in deaths and hospitalisations diverged from that in infection prevalence while the trend for hospitalisations started to reconverge during late April 2021 (Figure 9). When stratified by age, different patterns emerged. For deaths, continued divergence was observed at ages 65 years and above while the trends began to converge for those aged less than 65 years (Figure 10, Figure 11). However, rapid convergence between infection prevalence and hospital admissions was observed for the younger age group from May 2021 (Figure 10, Figure 11) — infections were substantially more frequent in the younger participants who were unvaccinated or who did not report vaccine status than in those who reported being vaccinated (Table 7).

**Table 7.**
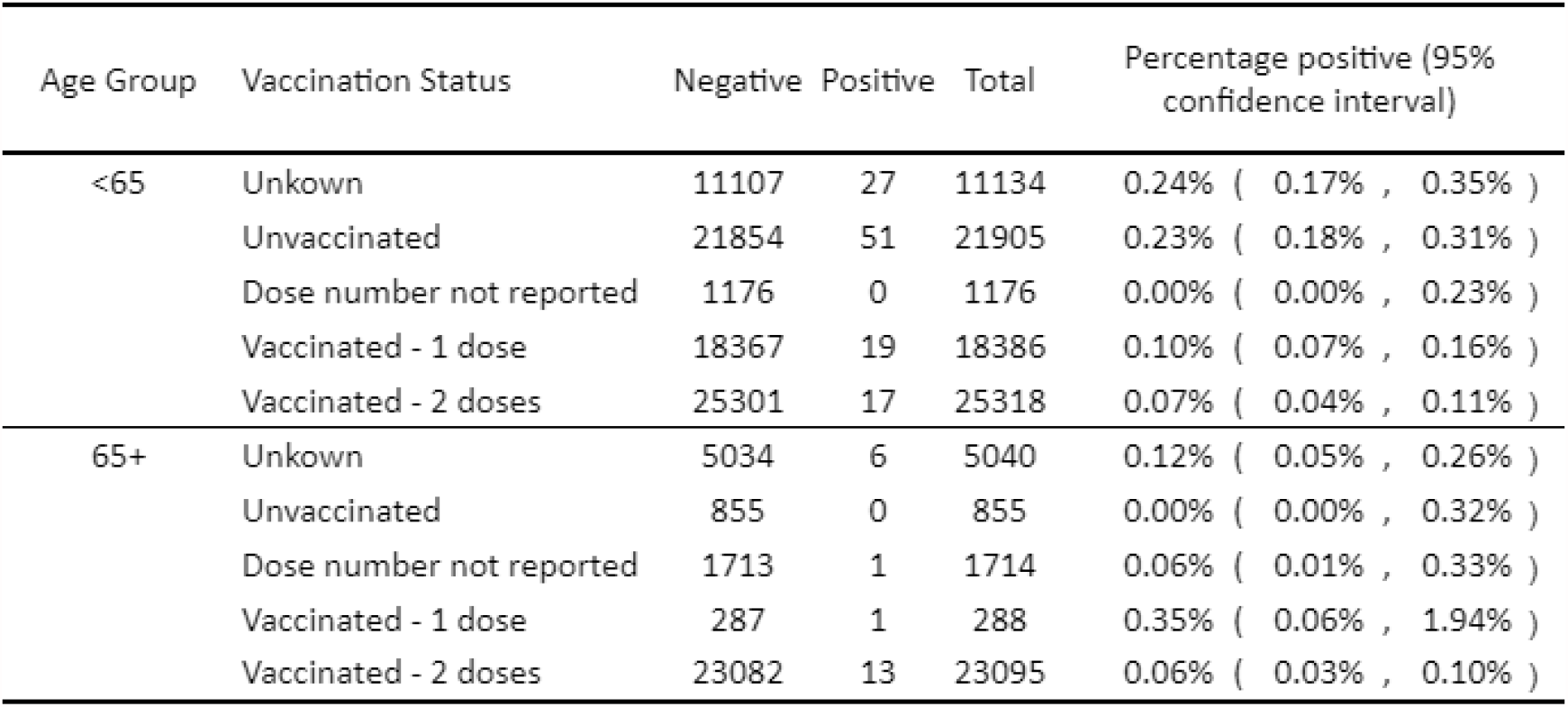
Prevalence of infection for younger and older age groups by self-reported vaccine status.

| Age Group | Vaccination Status | Negative | Positive | Total | Percentage positive (95% confidence interval) |
| --- | --- | --- | --- | --- | --- |
| <65 | Unkown | 11107 | 27 | 11134 | 0.24% ( 0.17% , 0.35% ) |
|  | Unvaccinated | 21854 | 51 | 21905 | 0.23% ( 0.18% , 0.31% ) |
|  | Dose number not reported | 1176 | 0 | 1176 | 0.00% ( 0.00% , 0.23% ) |
|  | Vaccinated - 1 dose | 18367 | 19 | 18386 | 0.10% ( 0.07% , 0.16% ) |
|  | Vaccinated - 2 doses | 25301 | 17 | 25318 | 0.07% ( 0.04% , 0.11% ) |
| 65+ | Unkown | 5034 | 6 | 5040 | 0.12% ( 0.05% , 0.26% ) |
|  | Unvaccinated | 855 | 0 | 855 | 0.00% ( 0.00% , 0.32% ) |
|  | Dose number not reported | 1713 | 1 | 1714 | 0.06% ( 0.01% , 0.33% ) |
|  | Vaccinated - 1 dose | 287 | 1 | 288 | 0.35% ( 0.06% , 1.94% ) |
|  | Vaccinated - 2 doses | 23082 | 13 | 23095 | 0.06% ( 0.03% , 0.10% ) |

**Figure 9.**
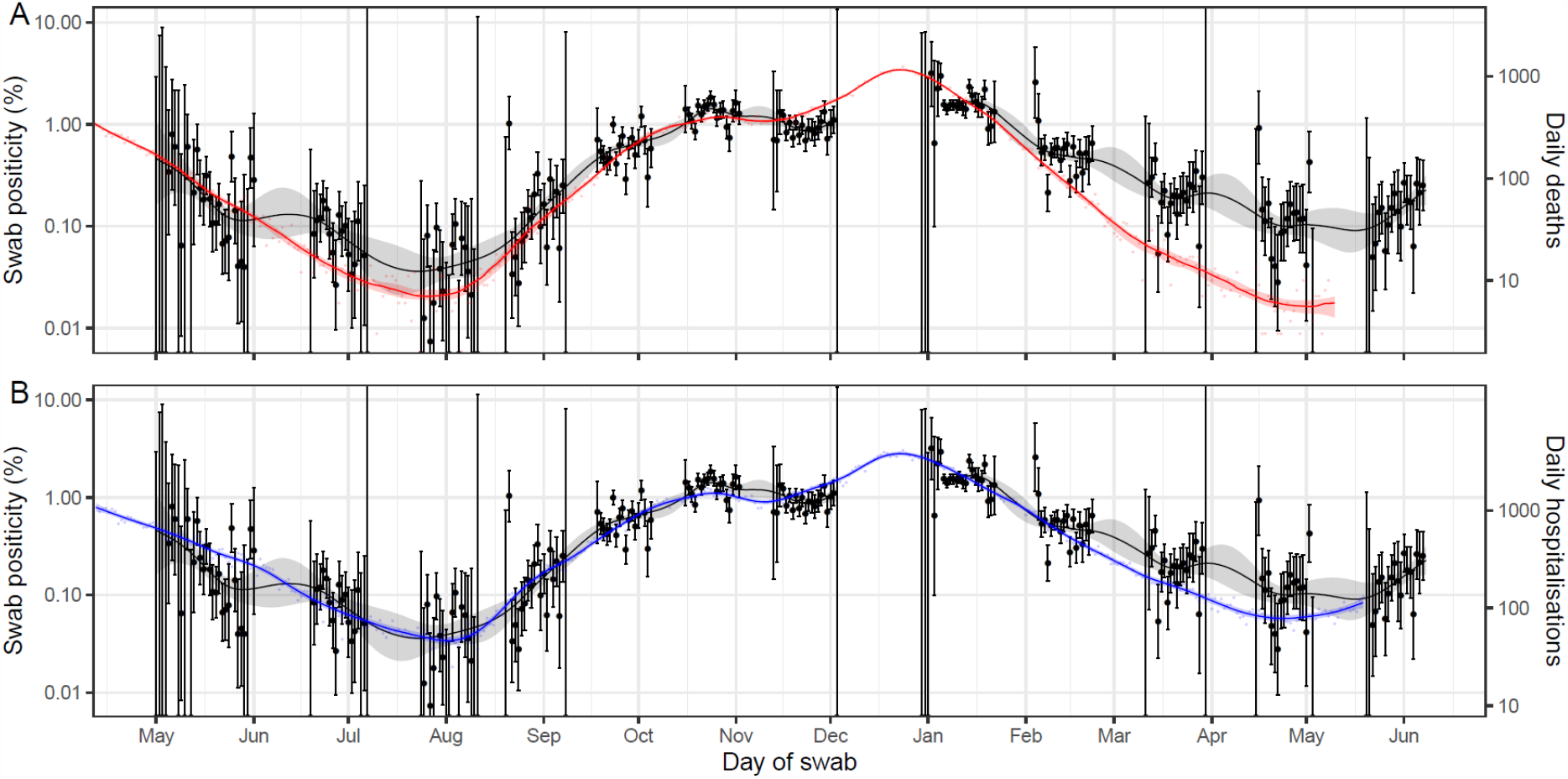
A comparison of daily deaths and hospitalisations to swab positivity as measured by REACT-1. Daily swab positivity for all 12 rounds of the REACT-1 study (black points with 95% confidence intervals, left hand y-axis) with P-spline estimates for swab positivity (solid black line, shaded area is 95% confidence interval). (A) Daily deaths in England (red points, right hand y-axis) and P-spline model estimates for expected daily deaths in England (solid red line, shaded area is 95% confidence interval, right hand y-axis). Daily deaths have been shifted by 26 (26, 26) days backwards in time along the x-axis. The two y-axis have been scaled using the best-fit scaling parameter 0.060 (0.059, 0.062). (B) Daily hospitalisations in England (blue points, right hand y-axis) and P-spline model estimates for expected daily hospitalisations in England (solid blue line, shaded area is 95% confidence interval, right hand y-axis). Daily hospitalisations have been shifted by 19 (19, 20) days backwards in time along the x-axis. The two y-axes have been scaled using the best-fit scaling parameter 0.24 (0.24, 0.25).

**Figure 10.**
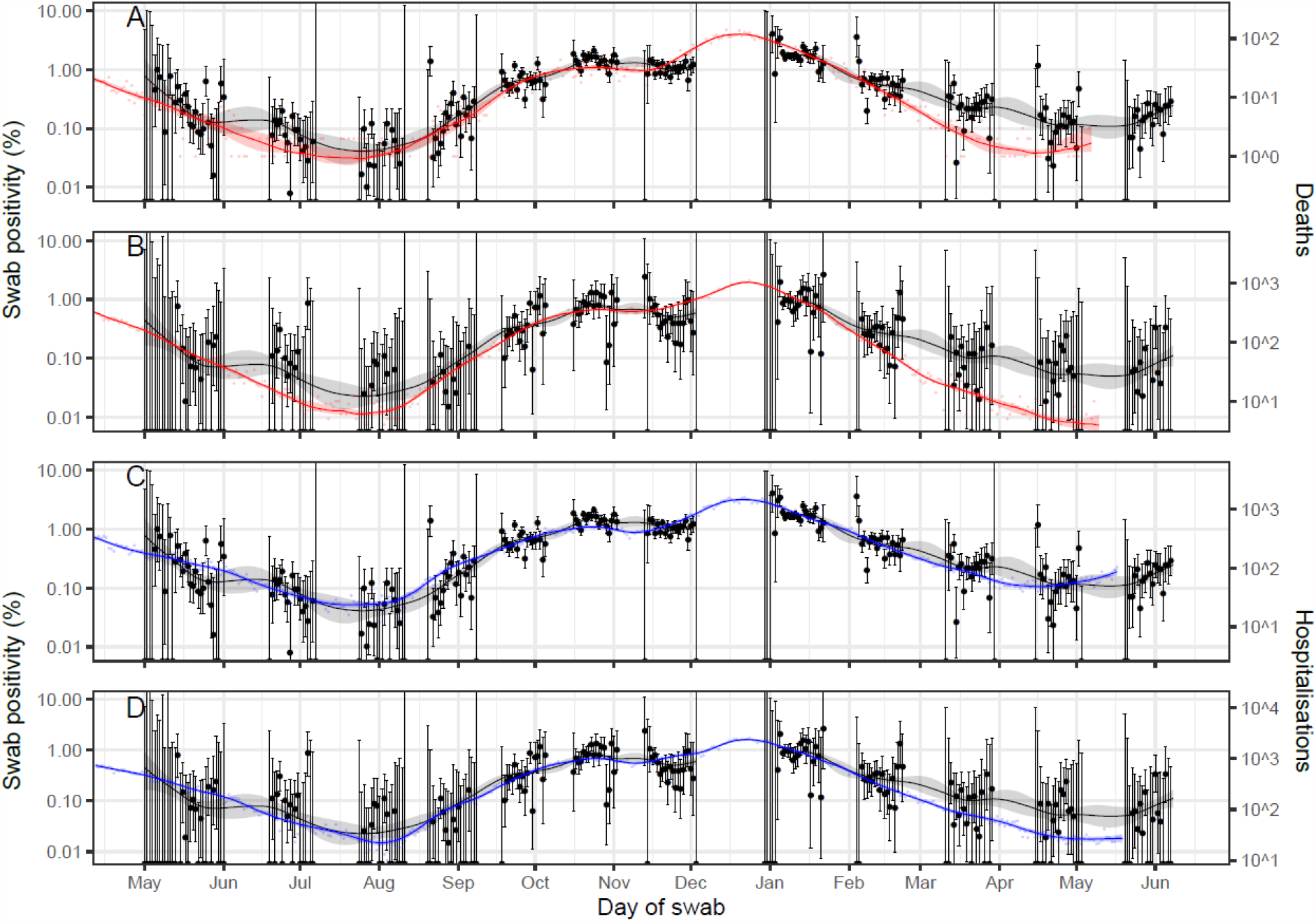
A comparison of daily deaths and hospitalisations to swab positivity as measured by REACT-1, by age group. Daily swab positivity for all 11 rounds of the REACT-1 study (black points with 95% confidence intervals, left hand y-axis) with P-spline estimates for swab positivity (solid black line, shaded area is 95% confidence interval) for (A, C) those aged under 64 and (B,D) those aged 65 and over. **(A)** Daily deaths for those aged 64 and under in England (red points, right hand y-axis) and corresponding P-spline model estimates for the expected number of deaths (solid red line, shaded area is 95% confidence interval, right hand y-axis). Daily deaths have been shifted by 29 (28, 29) days backwards in time along the x-axis. The two y-axis have been scaled using the best-fit scaling parameter 0.0065 (0.0063, 0.0066). **(B)** Daily deaths for those aged 65 and over in England (red points, right hand y-axis) and corresponding P-spline model estimates for the expected number of deaths (solid red line, shaded area is 95% confidence interval, right hand y-axis). Daily deaths have been shifted by 26 (24, 27) days backwards in time along the x-axis. The two y-axis have been scaled using the best-fit scaling parameter 0.52 (0.49, 0.55). **(C)** Daily hospitalisations for those aged 64 and under in England (blue points, right hand y-axis) and corresponding P-spline model estimates for the expected number of hospitalisations (solid blue line, shaded area is 95% confidence interval, right hand y-axis). Daily hospitalisations have been shifted by 21 (21, 21) days backwards in time along the x-axis. The two y-axis have been scaled using the best-fit scaling parameter 0.099 (0.097, 0.10). **(D)** Daily hospitalisations for those aged 65 and over in England (blue points, right hand y-axis) and corresponding P-spline model estimates for the expected number of hospitalisations (solid blue line, shaded area is 95% confidence interval, right hand y-axis). Daily hospitalisations have been shifted by 19 (17, 20) days backwards in time along the x-axis. The two y-axes have been scaled using the best-fit scaling parameter 1.4 (1.3, 1.5).

**Figure 11.**
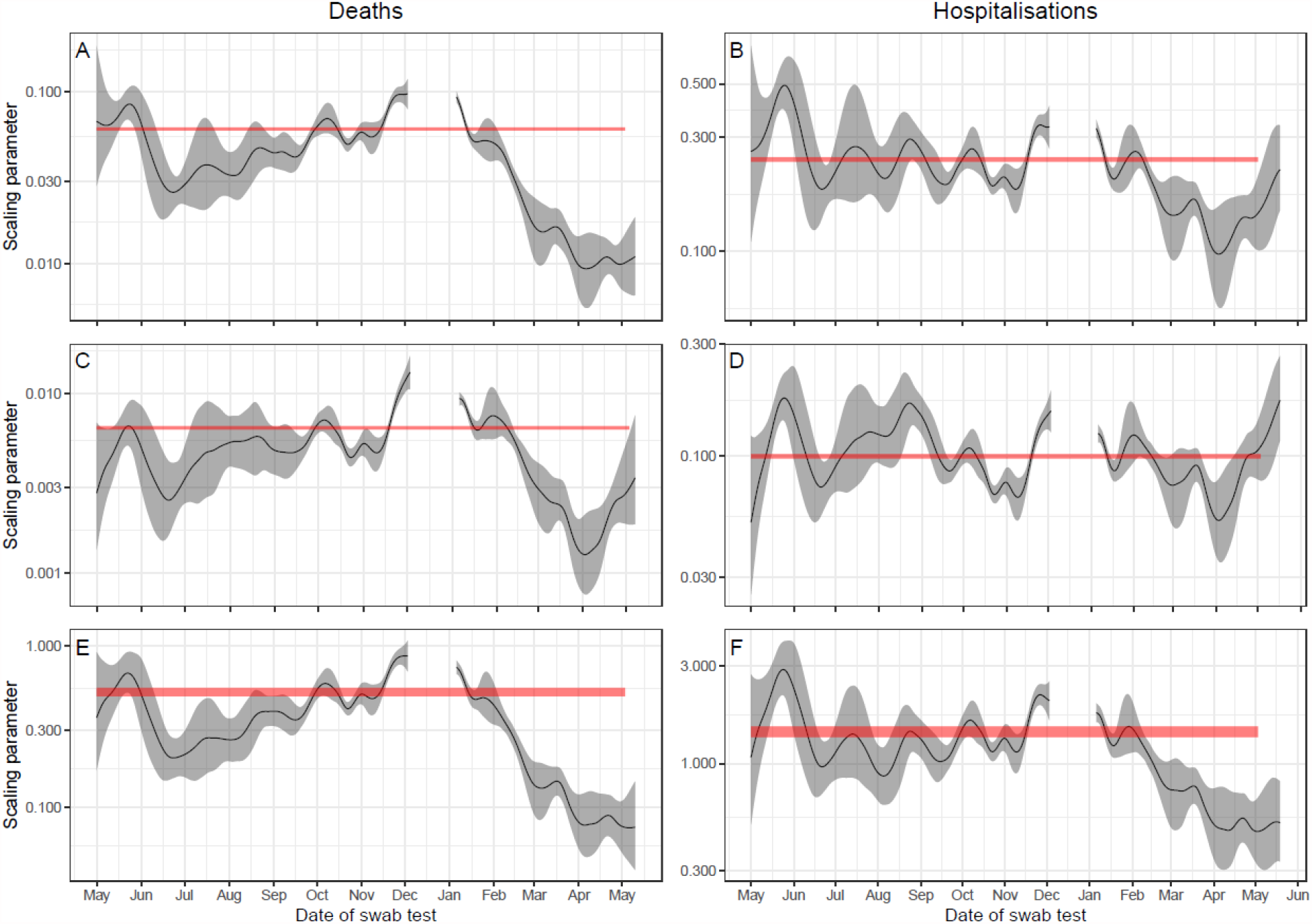
Comparison of REACT-1 swab-positivity P-spline estimates with time-lag adjusted P-spline estimates for death and hospitalisation data. Inferred scaling parameter (solid black line, grey shaded region is 95% confidence interval) directly calculated from the multiplicative difference between the REACT-1 P-splines for swab positivity and the death/hospitalisations P-splines, accounting for population size. 95% confidence intervals of the best-fitting average scaling parameters over the period of rounds 1 to 11 of the study (red shaded area) fit using the two parameter time-lag model are shown for comparison **(A)** Difference between REACT-1 swab positivity and deaths in all age groups assuming a time-lag of 26 (26, 26) days. **(B)** Difference between REACT-1 swab positivity and hospitalisations in all age groups assuming a time-lag of 19 (19, 20) days. **(C)** Difference between REACT-1 swab positivity and deaths in those aged 64 and under assuming a time-lag of 29 (28, 29) days. **(D)** Difference between REACT-1 swab positivity and hospitalisations in those aged 64 and under assuming a time-lag of 21 (21, 21) days. **(E)** Difference between REACT-1 swab positivity and deaths in those aged 65 and over assuming a time-lag of 26 (24, 27) days. **(F)** Difference between REACT-1 swab positivity and hospitalisations in those aged 65 and over assuming a time-lag of 19 (17, 20) days.

We observed the near replacement of the Alpha variant of SARS-CoV-2 with Delta during the period covered by rounds 11 and 12 of the study (15 April to 7 June) (Table 8). During round 12, the proportion of positive samples for which lineages could be obtained that were the Delta variant rose from ∼60% to ∼90% (Figure 12). Geographically, while we only detected Delta variant in two positive swabs in London during round 11 (15 April to 3 May), it became the sole lineage detected in the North West, East Midlands, South East and London during round 12 (Figure 13).

**Table 8.** Percentage of variants from positive samples for which a lineage could be accurately assessed.

| Round | N | Variant | n | Percentage |
| --- | --- | --- | --- | --- |
| 11 | 26 | Alpha | 24 | 92.3% ( 75.9% , 97.9% ) |
|  |  | Delta | 2 | 7.7% ( 2.1% , 24.1% ) |
| 12 | 46 | Alpha | 10 | 21.7% ( 12.3% , 35.6% ) |
|  |  | Delta | 36 | 78.3% ( 64.4% , 87.7% ) |

**Figure 12.**
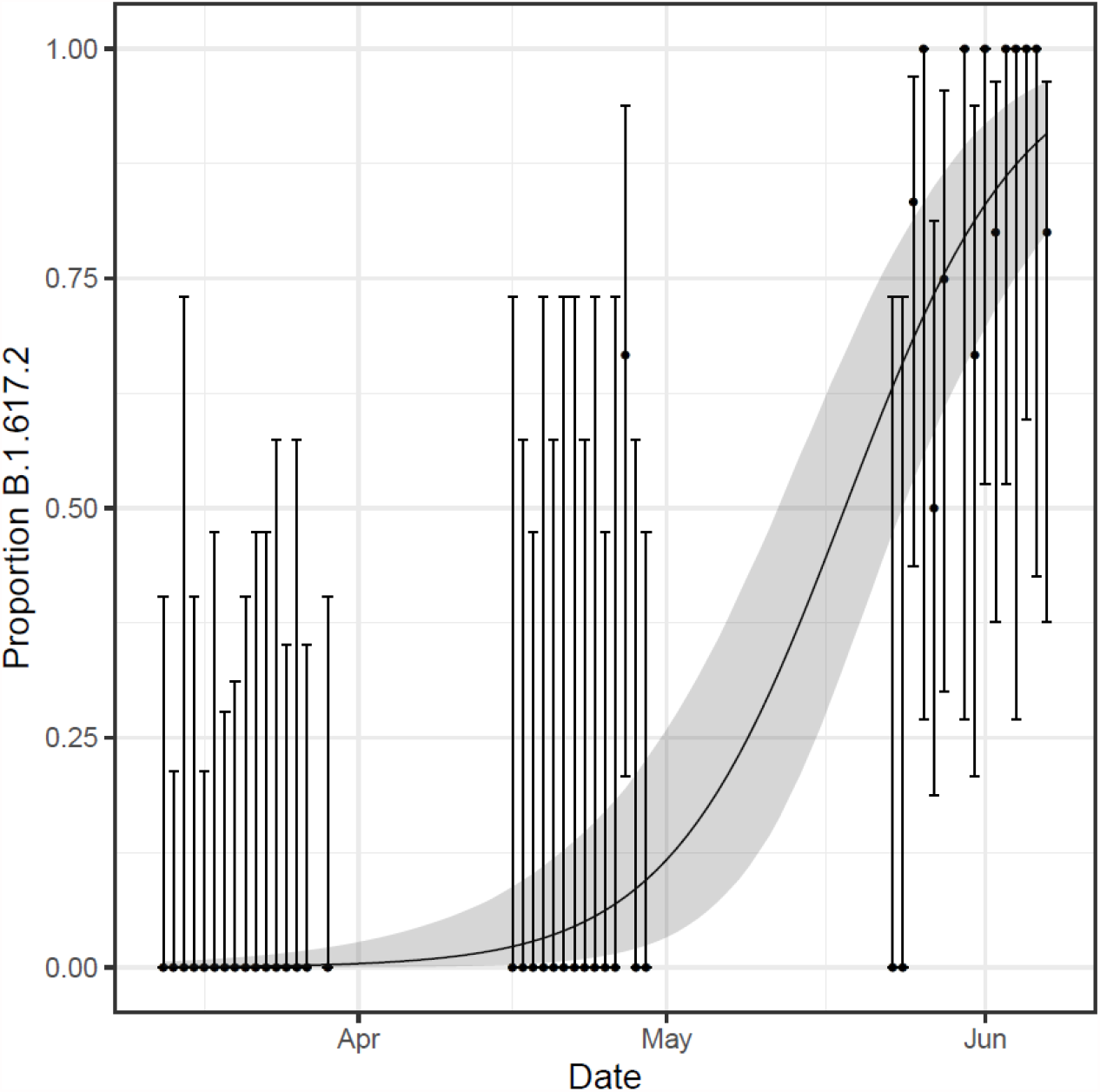
Plot of the proportion of Delta variant (B.1.617.2), among sequenced virus samples. Error bars show the 95% binomial confidence interval for each daily proportion calculation. Shaded region shows best-fit Bayesian logistic regression model with 95% credible interval.

**Figure 13.**
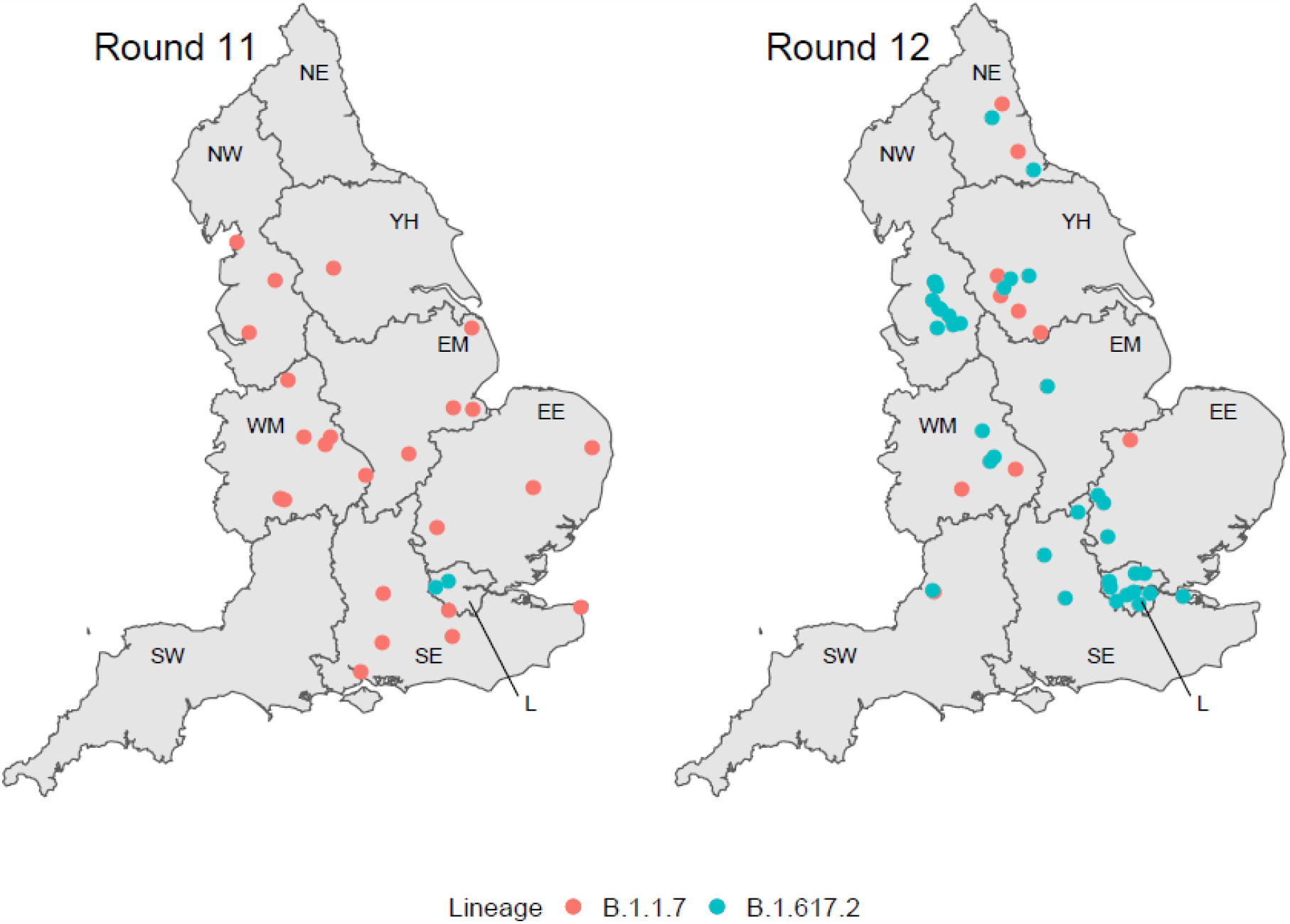
Jittered locations of positive samples for which sequencing could reliably determine lineages for rounds 11 (n=26) and 12 (n=46): B.1.1.7, Alpha; B.1.617.2, Delta. Regions: NE = North East, NW = North West, YH = Yorkshire and The Humber, EM = East Midlands, WM = West Midlands, EE = East of England, L = London, SE = South East, SW = South West.

## Discussion

Our results from REACT-1 round 12 showed an exponential increase in prevalence during the period 20 May to 7 June 2021, with prevalence rising 50% compared with the previous round (15 April to 7 May 2021). We estimate a doubling time of 11 days and R robustly above one at 1.44. This period of rapid growth coincides with the Delta variant becoming the dominant variant in England [13].

We observed that growth was being driven by younger age groups, with five-fold higher rates of swab-positivity among younger children (ages 5 to 12 years) and young adults (18 to 24 years) compared with those aged 65 years and older, and 2.5-fold higher rates among those below 50 years compared with those 50 years and above. These age patterns suggest that recent expansion of the vaccine programme to those aged 18 years and above [14] should help substantially to reduce the overall growth of the epidemic. The observed patterns may reflect increased social interactions among children and young adults as schools remain open and lockdown eases, as well as high vaccine uptake among older people.

Compared with our previous report covering the period 15 April to 3 May 2021, we have seen a rapid replacement of the Alpha variant by the Delta variant, such that at the beginning of the current round (20 May 2021) we estimate that around 60% of swab-positive tests were due to the Delta variant, rising to around 90% at the end of the round (7 June 2021).

We observed differing relationships between infection prevalence as measured in REACT-1 and hospitalisations and deaths from routine data sources; while we saw divergence of these patterns among the largely double-dose vaccinated population at older ages (65 years and above), at younger ages (less than 65 years) there has been a recent convergence in these patterns. This suggests a more direct link between infections and hospitalisations among this younger group with infections occurring predominantly among participants who were unvaccinated or did not provide a vaccination history. Our findings are in keeping with the high efficacy of the vaccines (ChAdOx1-S and BNT162b2) in preventing hospitalisations and deaths among the fully vaccinated population [15].

We found regional differences in swab-positivity with highest rates in the North West and lowest in the South West; though both northern and southern regions showed increasing trends in swab-positivity in the most recent data, compatible with the widespread distribution of the Delta variant across England. At a sub-regional level, there were pockets of high prevalence particularly in areas of the North West where surge testing is being undertaken in response to high infection rates [15,16].

Deprivation remained a key risk factor for swab-positivity, with a nearly two-fold higher adjusted odds of swab-positivity among participants living in the most deprived areas versus those in the least deprived areas. The finding of higher risk of infection among people living in the most deprived areas has been reported by us previously [17] and by others [18] likely reflecting a combination of working conditions, housing, overcrowding and access to outdoor space.

There are a number of limitations to our study. We changed our sampling strategy in round 12 in order to improve precision of prevalence estimates in more urban and deprived areas, especially at relatively low prevalence: rather than aiming to achieve similar sample sizes by lower-tier local authority (LTLA), as in previous rounds, we selected our sample to be proportionate to population by LTLA. This had the effect of increasing the numbers sampled in urban areas and decreasing those in rural areas. Although this may have affected comparison of unweighted prevalence across rounds, it should not affect comparisons of weighted estimates, since, in each round, we use weighting to correct our prevalence estimates to be representative of England as a whole.

Nonetheless, the change in sampling strategy will affect participation rates since participation is higher in more affluent rural areas (which have been down-sampled) than in more deprived urban areas (up-sampled). Here we report an overall response rate of 13.4% compared with 15.5% at the previous round [12]. Participation rates may also have been affected by the availability of ‘surge’ testing (including among non-symptomatic people) in areas of high prevalence. In addition, willingness to take part in a national surveillance programme such as REACT-1 may have reduced as lockdown eases and individuals are less likely to be available at home, e.g. for courier pick-up of the completed swab. Notwithstanding these limitations we believe that, by weighting by key socio-demographic variables, we are able to capture representative estimates of community prevalence of swab-positivity for SARS-CoV-2 over time, person and place across England. Furthermore, short-term trends in the data should be less affected by issues of non-response, allowing us to accurately characterise exponential growth.

In summary, we report exponential growth in prevalence of swab-positivity across England driven by younger age groups, during the period 20 May to 7 June. This was accompanied by a rapid replacement of the Alpha variant by the Delta variant. The extent to which exponential growth continues, or slows down as a consequence of the continued rapid roll-out of the vaccination programme, including to young adults, requires close monitoring. Data such as those presented here are vital to track the course of the epidemic and inform ongoing decisions about the timing of further lifting of restrictions in England.

## Methods

REACT-1 involves the collection of a self-administered throat and nose swab sample sent for RT-PCR testing from a random sample of the population in England at ages 5 years and above (administered by parent/guardian for ages 5 to 12 years) [10]. The population sample is obtained from the National Health Service (NHS) register of patients. Unlike previous rounds of REACT-1, we adjusted the sampling procedure in round 12 to select the sample randomly in proportion to population at LTLA level (previously we aimed for similar numbers of participants in each LTLA). To reflect the move into a lower prevalence regime, this revised sampling frame gives more weight to higher population density LTLAs in urban areas versus lower population density LTLAs in rural areas. However, data should be comparable across rounds as we re-weight the data to be representative of England as a whole. We sent out 814,633 invitations to people registered with a GP on the NHS register resulting in the dispatch of 161,037 (19.8%) test kits, yielding 108,911 (67.6%) completed swabs with a valid test result. This gave an overall response rate of 13.4% (valid swabs divided by total number of invitations).

We estimate RT-PCR swab-positivity both as crude prevalence without weighting, and to adjust for variable non-response we then weight the findings according to age, sex, local authority counts, ethnicity and deprivation. In this way, we aim to provide prevalence estimates by socio-demographic and other characteristics that are representative of the population of England as a whole.

We use exponential growth models to estimate the reproduction number R both across successive rounds and within rounds. We also provide estimates of R for i) different cut-points of cycle threshold (Ct) values for swab-positivity and ii) non-symptomatic people in the previous week.

We fit a smoothed P-spline function to the daily prevalence data (weighted) across all rounds, with knots at 5-day intervals, to investigate time-trends in prevalence, at all ages, for those under 50 years and those 50 years and above, and for Northern regions (including Midlands) and Southern regions. We then estimate scaling parameters corresponding to the percentage of people who are swab-positive in the population on a particular day that is associated with future hospitalisations or deaths offset by a suitable lag period — and compare daily prevalence data from rounds 1-11 of REACT-1 with publicly available national daily hospital admissions and COVID-19 mortality data (deaths within 28 days of a positive test). This gives an indication as to any alterations in the link between infection prevalence and subsequent hospitalisations and deaths, which we estimated both for all ages and for those aged under 65 years and 65 years and above.

Spatial analyses were undertaken based on nearest neighbours to estimate neighbourhood prevalence (the median number of neighbours within 20 km).

Viral genome sequencing was carried out for RT-PCR positive swab samples with sufficient sample volume and N-gene Ct values < 32. Viral RNA was amplified using the ARTIC protocol [16] with sequencing libraries prepared using CoronaHiT [19]. Sequencing data were analysed using the ARTIC bioinformatic pipeline [20] with lineages assigned using PangoLEARN [21].

Statistical analyses were carried out in R [14]. Research ethics approval was obtained from the South Central-Berkshire B Research Ethics Committee (IRAS ID: 283787). The COVID-19 Genomics UK Consortium (COG-UK) study protocol was approved by the Public Health England Research Ethics Governance Group (reference: R&D NR0195).

## Data Availability

The datasets generated or analysed, or both, during this study are not publicly available because of governance restrictions.

## Data availability

Supporting data for tables and figures are available either: in this [spreadsheet](https://docs.google.com/spreadsheets/d/18_GhlHxCuyF1yaWbo8wcAZb-FFlixrrGLiOxkjkxKi0/edit#gid=1057772495); or in the inst/extdata directory of this GitHub R package. Assembled/consensus genomes are available from GISAID subject to minimum quality control criteria. Raw reads are available from European Nucleotide Archive (ENA). All genomes, phylogenetic trees, and basic metadata are available from the COG-UK consortium website (https://www.cogconsortium.uk).

## Acknowledgements

SR, CAD acknowledge support: MRC Centre for Global Infectious Disease Analysis, National Institute for Health Research (NIHR) Health Protection Research Unit (HPRU), Wellcome Trust (200861/Z/16/Z, 200187/Z/15/Z), and Centres for Disease Control and Prevention (US, U01CK0005-01-02). NFA was supported by the Quadram Institute Bioscience BBSRC funded Core Capability Grant (project number BB/CCG1860/1). GC is supported by an NIHR Professorship. HW acknowledges support from an NIHR Senior Investigator Award and the Wellcome Trust (205456/Z/16/Z). PE is Director of the MRC Centre for Environment and Health (MR/L01341X/1, MR/S019669/1). PE acknowledges support from Health Data Research UK (HDR UK); the NIHR Imperial Biomedical Research Centre; NIHR HPRUs in Chemical and Radiation Threats and Hazards, and Environmental Exposures and Health; the British Heart Foundation Centre for Research Excellence at Imperial College London (RE/18/4/34215); and the UK Dementia Research Institute at Imperial (MC_PC_17114). We thank The Huo Family Foundation for their support of our work on COVID-19. Quadram authors gratefully acknowledge the support of the Biotechnology and Biological Sciences Research Council (BBSRC); their research was funded by the BBSRC Institute Strategic Programme Microbes in the Food Chain BB/R012504/1 and its constituent project BBS/E/F/000PR10352. We thank members of the COVID-19 Genomics Consortium UK for their contributions to generating the genomic data used in this study. The COVID-19 Genomics UK (COG-UK) Consortium is supported by funding from the Medical Research Council (MRC) part of UK Research & Innovation (UKRI), the National Institute of Health Research (NIHR) and Genome Research Limited, operating as the Wellcome Sanger Institute.

We thank key collaborators on this work – Ipsos MORI: Kelly Beaver, Sam Clemens, Gary Welch, Nicholas Gilby, Kelly Ward and Kevin Pickering; Institute of Global Health Innovation at Imperial College: Gianluca Fontana, Sutha Satkunarajah, Didi Thompson and Lenny Naar; Molecular Diagnostic Unit, Imperial College London: Prof. Graham Taylor; North West London Pathology and Public Health England for help in calibration of the laboratory analyses; Patient Experience Research Centre at Imperial College and the REACT Public Advisory Panel; NHS Digital for access to the NHS register; and the Department of Health and Social Care for logistic support. SR acknowledges helpful discussion with attendees of meetings of the UK Government Office for Science (GO-Science) Scientific Pandemic Influenza – Modelling (SPI-M) committee.

## Additional information

Full list of COG-UK author’s names and affiliations are available in this [spreadsheet](https://drive.google.com/file/d/11Q3cnp2qGKMqcfrTKpp53jZFc7OdF0f7/view).

